# A Multi-Context Regulome-Wide Association Atlas for Genetic Studies of Aging Brain Disorders

**DOI:** 10.64898/2026.05.15.26353329

**Authors:** Chunming Liu, Anqi Wang, Hao Sun, Kaixuan Luo, Sheng Qian, Yining Li, Daniel Nachun, Xin He, Philip De Jager, David Bennett, Minghui Wang, Carlos Cruchaga, The Alzheimer’s Disease Functional Genomics Consortium, Gao Wang, Fabio Morgante

## Abstract

Genome-wide association studies have identified risk loci for aging brain disorders, but mechanistic interpretation depends on linking these loci to genes and to the tissues, cell types, and molecular modalities in which they act. Here, we introduce FunGen-xQTL Multi-Brain (FGMB), a multi-context atlas of *cis*-regulatory genetic prediction models for regulome-wide association studies (RWAS), built from molecular datasets assembled by the ADSP Functional Genomics Consortium (FunGen-AD). FGMB provides *cis*-genetic prediction models for 17,901 protein-coding genes across 36 molecular datasets, 18 contexts, and 3 regulatory modalities, comprising more than 293,000 imputable gene-level and splice-event models. These models are derived from eight sparse regression, Bayesian and multivariate prediction methods, including cross-context approaches that integrate information across tissues and cell types. We applied FGMB to Alzheimer’s disease and identified 327 RWAS associations. Multi-context joint variant–gene fine-mapping then resolved 86 gene–molecular-trait pairs as regulatory signals rather than linkage disequilibrium (LD) hitchhiking.

## INTRODUCTION

Genome-wide association studies (GWAS) have mapped many loci for complex traits, including aging brain disorders, but most associated variants lie in non-coding regions and do not directly identify target genes or regulatory contexts^1,2^. Because genetic regulation is often tissue-, cell-type-, or modality-specific, translating GWAS loci into mechanism depends on connecting variants to genes and to the functional contexts in which those genes are regulated^3–5^. Transcriptome-wide association studies (TWAS), and regulome-wide association studies (RWAS) more generally, provide one such connection by testing whether genetically predicted molecular traits are associated with disease risk^6,7^. Public TWAS weight repositories, including PredictDB/*MetaXcan* and FUSION, have made this strategy broadly reusable through GTEx-derived bulk-tissue gene expression models, with PredictDB/*MetaXcan* also providing large GTEx v8 splicing prediction models^3,6–8^. Additional brain-relevant prediction resources with downloadable model files have been released for selected settings, including PsychENCODE cross-disorder adult-cortex models, BrainSeq Phase II dorsolateral prefrontal cortex and hippocampus models, and multi-region Alzheimer’s disease (AD) neocortical transcriptome models^9–11^. In parallel, large brain and AD functional-genomic studies, including CommonMind, MetaBrain, human microglia, and single-nucleus brain eQTL resources, provide rich xQTL maps and molecular annotations, yet many distribute QTL summary statistics, browsers, or controlled-access molecular data rather than standardized RWAS prediction weights^4,5,12,13^. A resource for aging brain disorders therefore needs a harmonized framework that represents postmortem brain datasets, brain regions, single-nucleus cell types and subtypes, peripheral immune contexts, and regulatory modalities beyond RNA abundance, including protein abundance and splicing.

A useful RWAS resource is defined not only by the molecular datasets it includes, but also by the statistical models used to train and select prediction weights. Many standard RWAS resources still rely on sparse-regression predictors, including *elastic-net* and *lasso*^6,7^. These models are useful predictors, but they can be restrictive when regulatory effects are distributed across variants, shared across contexts, or estimated from reference panels with limited effective sample size^14,15^. Bayesian shrinkage and fine-mapping models can accommodate a wider range of genetic architectures. Multi-tissue or multi-context models can also borrow information across related tissues or cell types to improve prediction accuracy and distinguish context-shared from context-specific regulation^15–17^. These developments motivate a RWAS resource that is broad both biologically and statistically, including diverse molecular traits and prediction models.

Even with accurate predictors, marginal RWAS associations require refinement before they can be interpreted as *candidate* regulatory mechanisms. Gusev *et al.* introduced joint and conditional TWAS analyses to distinguish multiple marginally associated genes at a locus by evaluating association signals after conditioning on all those genes^7^. Later work showed that marginal TWAS can nominate genes whose predictors are correlated with nearby variants or with other genetically regulated molecular traits, producing false discoveries through linkage disequilibrium (LD) hitchhiking or co-regulation^18,19^. Recent analyses further showed that predictors from non-trait-relevant tissues can tag GWAS-associated haplotypes through tightly linked regulatory variants, making TWAS associations unreliable^20^. This issue becomes more significant in multi-molecular-trait resources, where predicted molecular traits are correlated across tissues, cell types, modalities, and genes^21^. An aging brain regulome-wide association atlas should therefore couple broad predictor construction with joint variant–gene fine-mapping across molecular traits, so that tissue- and cell-type-specific associations can be evaluated against cross-molecular-trait tagging.

Here, we introduce FunGen-xQTL Multi-Brain (FGMB), a multi-context regulome-wide association atlas for genetic studies of aging brain disorders. FGMB integrates molecular datasets assembled by FunGen-AD, spanning brain regions, single-nucleus cell types and subtypes, peripheral monocytes, gene expression, protein abundance, and splicing. Across 36 molecular datasets, 18 contexts, and 3 regulatory modalities, FGMB provides *cis*-genetic prediction models for approximately 17,901 protein-coding genes, yielding more than 293,000 imputable gene-level or splice-event models. For each molecular dataset, FGMB evaluates eight prediction models, including established TWAS sparse-regression methods (*lasso* and *elastic-net*), Bayesian methods less commonly used in RWAS weight resources (*BayesL*, *BayesR*, *mr.ash*, and *SuSiE*), and multivariate cross-context methods (*mr.mash* and *mvSuSiE*) that learn shared regulatory effects across molecular traits. As an illustration, we applied 11 ROSMAP molecular datasets to AD GWAS summary statistics^22^, identified 327 RWAS associations corresponding to 138 unique genes. We also used multi-context joint variant–gene fine-mapping to prioritize 86 RWAS-significant gene–molecular-trait pairs with suggestive evidence of non-zero effects. FGMB is released as part of the FunGen-AD resource, with molecular-trait-specific predictors and downstream analysis results. By pairing broad molecular scale with diverse model selection and joint variant–gene fine-mapping, FGMB is designed to expand genetic regulation of aging brain disorders, while distinguishing predicted molecular-trait associations from variant-driven signals.

## RESULTS

FGMB combines three linked components in one resource-to-application workflow. First, it builds a molecular prediction resource spanning 36 molecular datasets, 18 contexts, and 3 regulatory modalities, using Bayesian and cross-context model selection to improve RWAS weights. Second, it applies the selected weights to AD GWAS summary statistics to obtain RWAS associations. Third, it performs joint variant–gene fine-mapping to distinguish associations with evidence of non-zero effect from those due to LD hitchhiking and co-regulation, thereby enhancing locus-level interpretation. The atlas is organized for reuse through a The National Institute on Aging Genetics of Alzheimer’s Disease Data Storage Site (NIAGADS xQTL) portal landing page that links FGMB prediction weights, association results, fine-mapping outputs, LD reference panel, and step-by-step RWAS vignettes.

### FGMB Provides Molecular Trait Prediction Models Across Aging-Brain Contexts

We constructed *cis*-genetic prediction models for gene expression, protein abundance, and splicing regulation across molecular datasets assembled by FunGen-AD, spanning ROSMAP^23^, MSBB^24^, and Knight-ADRC^25^. FGMB includes bulk brain regions, peripheral monocytes, single-nucleus cell types and subtypes, and three molecular modalities (*i.e.*, gene expression, protein abundance, and splicing regulation). In total, the release spans 36 molecular datasets, comprising 18 contexts, and 3 regulatory modalities and yielding more than 293,000 imputable gene-level or splice-event models. The single-nucleus component includes CUIMC1, MIT, and a CUIMC1/CUIMC2/MIT mega-analysis for six major cell types; CUIMC2 is a recently generated ROSMAP DLPFC single-nucleus multiome (RNA-seq + ATAC-seq) dataset from approximately 240 individuals^26^. Dataset-level composition, sample sizes, and definitions are provided in Supplementary Note S1 and summarized in Table 1.

**Table 1.** Summary of FGMB Molecular Trait Prediction Models. *Molecular Dataset* refers to a source-specific molecular measurement panel used for predictor training, such as a brain region, cell type, or cell subtype measured for gene expression, protein abundance, or splicing. *Context*denotes the tissue, cell type, or cell subtype, whereas *Molecular Modality* denotes the type of molecular phenotype being modeled.

| Source Dataset | Molecular Dataset | Context | Molecular Modality | Sample Size | Genes Trained | Imputable Genes |
| --- | --- | --- | --- | --- | --- | --- |
| ROSMAP<sup>23</sup> | DLPFC eQTL | Dorsolateral Prefrontal Cortex | Gene Expression | 777 | 16307 | 9948 |
|  | PCC eQTL | Posterior Cingulate Cortex | Gene Expression | 441 | 16110 | 10720 |
|  | AC eQTL | Anterior Cingulate Cortex | Gene Expression | 593 | 16104 | 10918 |
|  | Mono eQTL | Monocyte | Gene Expression | 226 | 12801 | 5397 |
| ROSMAP<sup>27</sup> | DLPFC pQTL | Dorsolateral Prefrontal Cortex | Protein Expression | 416 | 7396 | 3925 |
| ROSMAP<sup>28</sup> | DLPFC sQTL | Dorsolateral Prefrontal Cortex | Splicing Enrichment | 806 | 12474 | 7261 |
|  | PCC sQTL | Posterior Cingulate Cortex | Splicing Enrichment | 449 | 12663 | 9832 |
|  | AC sQTL | Anterior Cingulate Cortex | Splicing Enrichment | 603 | 12585 | 8472 |
| ROSMAP<sup>5</sup> | Ast eQTL CUIMC1 | Astrocyte | Gene Expression | 419 | 11392 | 6023 |
|  | Inh eQTL CUIMC1 | Inhibitory Neuron | Gene Expression | 419 | 11266 | 6609 |
|  | Exc eQTL CUIMC1 | Excitatory Neuron | Gene Expression | 419 | 11111 | 8085 |
|  | Oli eQTL CUIMC1 | Oligodendrocyte | Gene Expression | 419 | 10912 | 5595 |
|  | OPC eQTL CUIMC1 | Oligodendrocyte Progenitor | Gene Expression | 418 | 7742 | 3582 |
|  | Mic eQTL CUIMC1 | Microglia | Gene Expression | 419 | 7130 | 3071 |
| ROSMAP<sup>29</sup> | Ast eQTL MIT | Astrocyte | Gene Expression | 385 | 9125 | 5530 |
|  | Ast.10 eQTL | Astrocyte Subtype 10 | Gene Expression | 113 | 1927 | 1927 |
|  | Inh eQTL MIT | Inhibitory Neuron | Gene Expression | 379 | 10760 | 7141 |
|  | Exc eQTL MIT | Excitatory Neuron | Gene Expression | 386 | 10645 | 8000 |
|  | Oli eQTL MIT | Oligodendrocyte | Gene Expression | 387 | 10021 | 6748 |
|  | OPC eQTL MIT | Oligodendrocyte Progenitor | Gene Expression | 383 | 8707 | 5163 |
|  | Mic eQTL MIT | Microglia | Gene Expression | 377 | 5404 | 3306 |
|  | Mic.12 eQTL MIT | Microglia Subtype 12 | Gene Expression | 106 | 702 | 701 |
|  | Mic.13 eQTL MIT | Microglia Subtype 13 | Gene Expression | 80 | 692 | 692 |
| ROSMAP<sup>26</sup> | Ast eQTL mega | Astrocyte | Gene Expression | 737 | 7742 | 4428 |
|  | Inh eQTL mega | Inhibitory Neuron | Gene Expression | 736 | 9577 | 5970 |
|  | Exc eQTL mega | Excitatory Neuron | Gene Expression | 737 | 10138 | 7856 |
|  | Oli eQTL mega | Oligodendrocyte | Gene Expression | 737 | 8196 | 4826 |
|  | OPC eQTL mega | Oligodendrocyte Progenitor | Gene Expression | 735 | 5897 | 2787 |
|  | Mic eQTL mega | Microglia | Gene Expression | 733 | 3514 | 1562 |
| MSBB<sup>24</sup> | FP eQTL | Frontal Pole | Gene Expression | 274 | 9275 | 8088 |
|  | STG eQTL | Superior Temporal Gyrus | Gene Expression | 254 | 9275 | 7645 |
|  | PHG eQTL | Parahippocampal Gyrus | Gene Expression | 230 | 9275 | 7494 |
|  | IFG eQTL | Inferior Frontal Gyrus | Gene Expression | 256 | 9275 | 7997 |
|  | PHG pQTL | Parahippocampal Gyrus | Protein Expression | 184 | 11224 | 8325 |
| Knight-ADRC<sup>25</sup> | PC eQTL | Parietal Cortex | Gene Expression | 354 | 15941 | 9070 |
|  | PC pQTL | Parietal Cortex | Protein Expression | 412 | 1018 | 233 |

### Bayesian and Cross-Context Models Improve Molecular Trait Prediction

We assessed prediction performance in the 11 ROSMAP molecular datasets used for the primary AD application, spanning bulk-tissue expression, monocyte expression, DLPFC protein abundance, and CUIMC1 single-nucleus expression. Across these datasets, we obtained predictors for 16,976 genes. Imputability was evaluated among gene–molecular-trait pairs for which at least one training method produced an imputable predictor, defined as cross-validated *R*^2^ *≥* 0.01 and *P <* 0.05. In total, 73,874 unique gene–molecular-trait pairs were imputable, corresponding to 15,423 unique genes (Table 1), among 128,290 total trained gene–molecular-trait pairs. On average, 57.7% of gene–molecular-trait pairs yielded imputable models from at least one prediction method (Figure 1A). Across all prediction models for imputable gene–molecular-trait pairs, the median cross-validated prediction accuracy was *R*^2^ = 0.023, which increased to *R*^2^ = 0.044 among the prediction models selected for the RWAS association analysis. Bulk RNA-seq contexts (*i.e.*, AC, PCC, and DLPFC) generally showed more imputable genes and stronger prediction performance than cell-type contexts (*i.e.*, Mono, Ast, and Inh) (Figure 1B). Bulk tissue gene expression datasets yielded 61.0%–67.8% of genes as imputable by at least one method, whereas cell-type molecular datasets ranged from 42.2%–72.8%. Imputability for cell-type molecular datasets depends on cell-type proportion. For example, Exc reached 72.8% imputability, approaching the bulk-tissue range. In fact, in the CUIMC1 resource, Exc constituted the largest cell-type proportion across donors, with a median of 1,618 cells per donor (43%), compared with 735 (20%) for Oli and 561 (15%) for Inh^5^.

**Figure 1.**
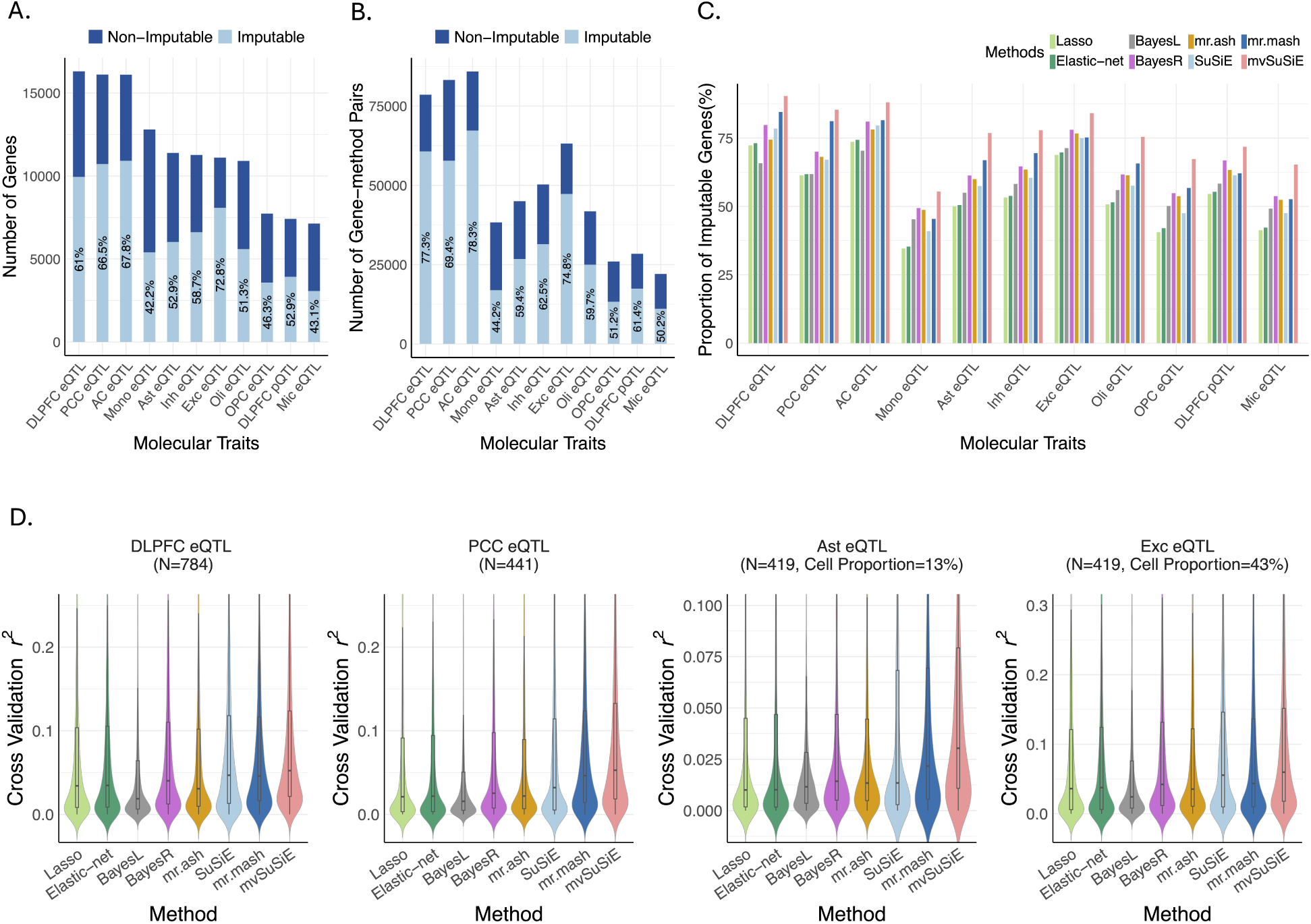
FGMB Molecular Trait Prediction Model Coverage and Performance. (A) Number of total gene–molecular-trait pairs and the proportion of imputable gene–molecular-trait pairs for each molecular trait. Imputable gene–molecular-trait pairs were identified when cross-validation *R*^2^ ≥ 0.01 and *p <* 0.05 from at least one of the eight methods. (B) Number of total gene–molecular-trait-method pairs for gene–molecular-trait pairs with an imputable prediction model from at least one method. (C) Proportion of imputable gene–molecular-trait pairs by each method. (D) Prediction model cross-validation *R*^2^ performance across molecular traits.

Prediction performance varied by model. The *elastic-net* and *lasso* reached 57.8% and 56.9% imputability, respectively. Bayesian and multivariate methods showed higher imputability, with *BayesL* (60.8%), *BayesR* (68.9%), *mr.ash* (66.8%), *SuSiE* (62.7%), *mr.mash* (72.8%), and *mvSuSiE* (80.5%). Relative to a two-method benchmark based on *elastic-net* and *lasso*, the full eight-method panel added 28,014 imputable gene–molecular-trait pairs, corresponding to 7,327 additional unique genes, which correspond to 37.9% of the final imputable set in the primary ROSMAP AD application datasets. Prediction accuracy also improved when the final RWAS weights were selected from the full model panel, with median cross-validation *R*^2^ increasing from 0.0203 under the two sparse-regression models to 0.0438 (a 115.8% relative improvement) after selecting the best-performing model from all eight methods. The models selected from the two multivariate Bayesian methods, *mr.mash* and *mvSuSiE*, reached a median *R*^2^ of 0.0455, corresponding to a 124.3% relative improvement over the models selected by *elastic-net* and *lasso*. Notably, the two multivariate Bayesian methods yielded imputable predictors for 9,410 gene–molecular-trait pairs that were non-imputable using any univariate approach. These gains support the use of Bayesian and multivariate methods as a core component of aging-brain RWAS weight construction. Additional context-specific performance patterns are provided in Supplementary Note S2. The methods and pipelines to compute RWAS weights have been implemented in the `xqtl-protocol` resource and the `pecotmr` R package, which makes the FGMB weight-building workflow reusable with additional molecular datasets, increasing the reach of the work.

Beyond the 73,874 imputable gene–molecular-trait pairs in the primary ROSMAP AD application datasets, the same model-building framework produced 219,969 additional imputable gene-level or splice-event models across ROSMAP bulk splicing, additional ROSMAP single-nucleus resources, MSBB, and Knight-ADRC (Supplementary Figures S1–S5; Supplementary Note S1). These analyses indicate that the improvements from Bayesian and multivariate models was not restricted to the primary AD application datasets, supporting FGMB as a scalable aging-brain RWAS weight and protocol resource.

### Alzheimer’s Disease RWAS Identifies Context-Level Gene–Molecular-Trait Associations

We next applied the ROSMAP component of FGMB to AD GWAS summary statistics. Using imputable molecular trait predictors across 11 ROSMAP molecular traits and summary statistics from Bellenguez *et al.*^22^, we tested 15,423 unique genes and identified 327 gene–molecular-trait pairs with significant RWAS associations at a Bonferroni-corrected threshold of *p <* 6.77 *×* 10^−7^ (obtained as 0.05/73,874, where the denominator is the number of imputable gene–molecular-trait pairs) (Figure 2A), corresponding to 138 unique genes. Among these 138 genes, 48 (34.8%) colocalized with AD GWAS signals^30^. The significant genes included well-established AD risk genes such as *BIN1*, *CR1*, *CLU*, *PICALM*, *APP*, and *APOE*. We also found 42 genes with previously unreported associations with AD risk (Table 2), including 9 gene–molecular-trait associations identified exclusively by multivariate methods. Several of the strongest previously unreported associations were also RWAS-significant using three additional AD GWAS studies, including Kunkle *et al.*^31^, Jansen *et al.*^32^, and Wightman *et al.*^33^. These included *IRF2BP1* -DLPFC pQTL, *HIF3A*-PCC eQTL, *ZNF576* -Ast eQTL, *ZNF428* -Oli eQTL, and *FBXO46* -Ast eQTL (Table 2). Supplementary Table S3 provides cross-GWAS replication across all four AD GWAS summary statistics, and Supplementary Table S1 provides molecular-trait-specific RWAS association results. The complete RWAS association results are available through the Synapse portal (see Data Availability).

**Figure 2.**
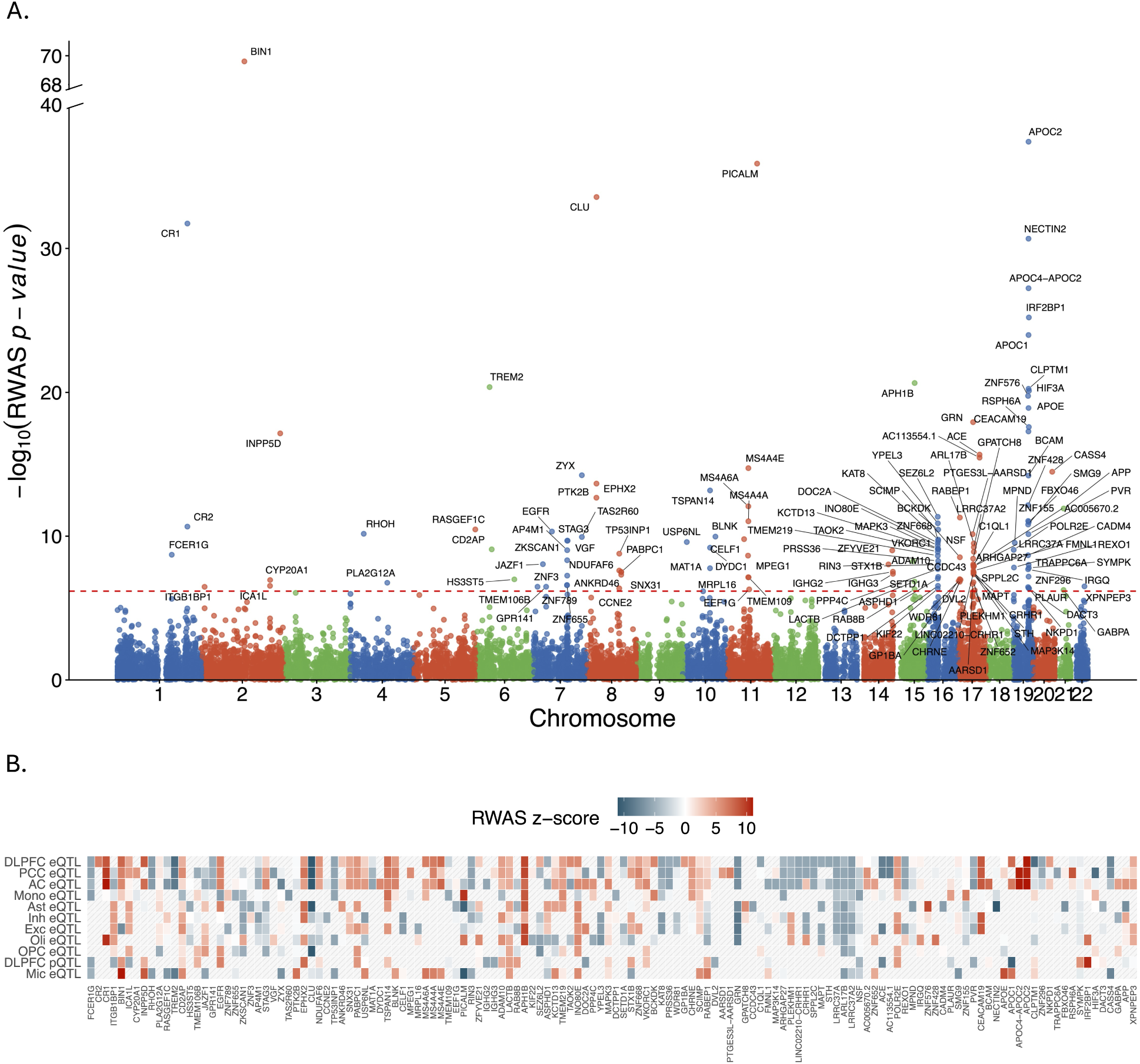
Alzheimer’s Disease RWAS Association Landscape Across Molecular Traits. (A) Manhattan plot for RWAS associations among all molecular traits, showing the most significant RWAS *p*-value for each gene across all tested molecular traits, providing a genome-wide view of association strength.(B) Heatmap of RWAS associations for genes with significant associations in at least one molecular trait based on RWAS *p*-value, with genes ordered by chromosomal position and molecular traits shown in rows. Positive *z*-scores (red) indicate that higher predicted molecular trait values are associated with increased AD risk, whereas negative *z*-scores (blue) indicate that higher predicted molecular trait values are associated with decreased risk (potentially protective effects). Striped grey boxes indicate gene–molecular-trait pairs with missing molecular data or prediction models that failed to meet imputability criteria (cross-validation *R*^2^ *<* 0.01 or *p >* 0.05).

**Table 2.**
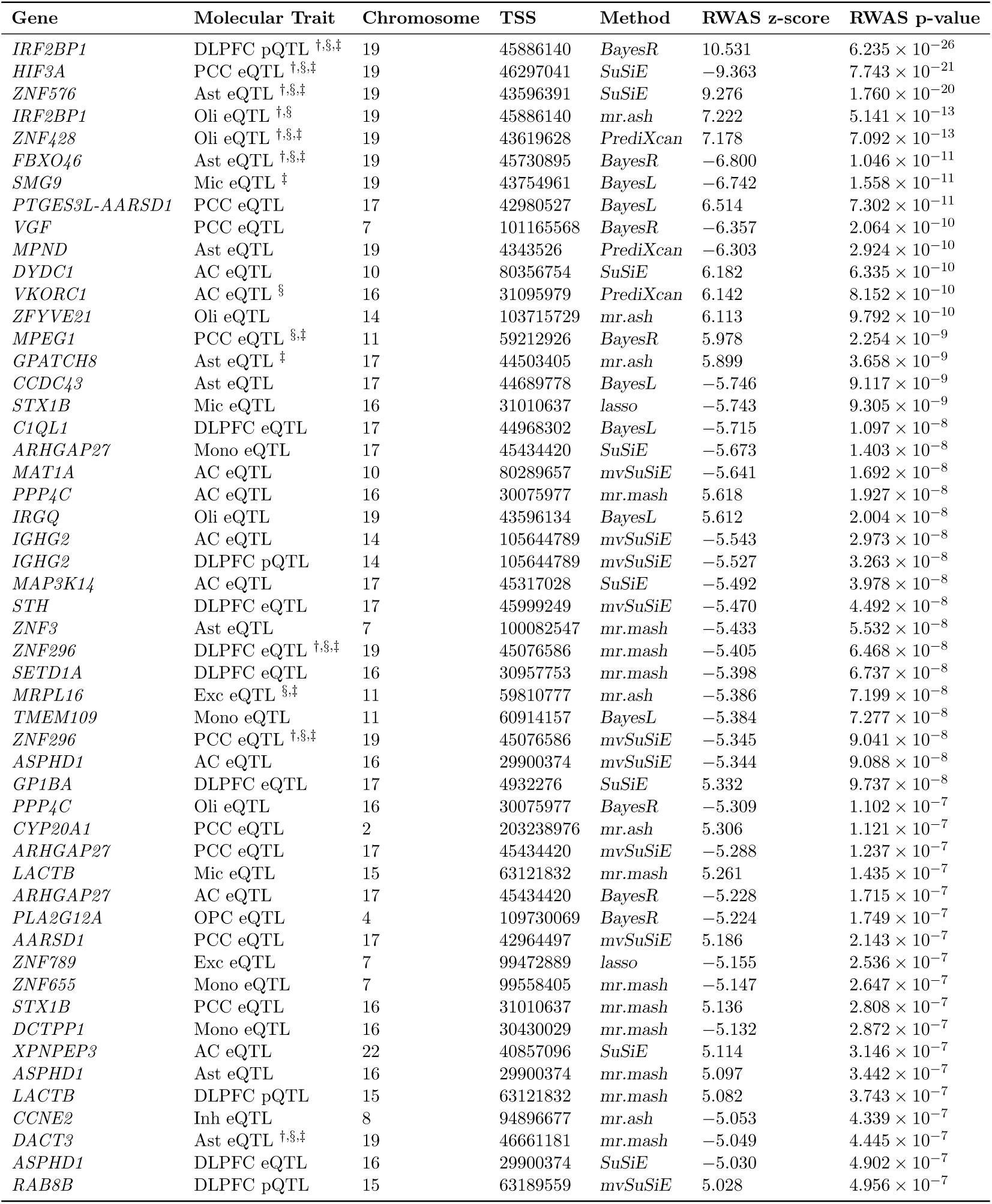
Summary of Previously Unreported RWAS Association Results. *Method* denotes the model used to train molecular trait predictors (*e.g.*, *lasso*, *elastic-net*). Only genes surpassing the Bonferroni-corrected significance threshold (*p <* 6.77 × 10^−7^) are shown. RWAS was performed using GWAS summary statistics from Bellenguez *et al.*. Previously unreported genes are defined as RWAS-significant genes not previously reported by large-scale GWAS, RWAS, or curated gene resources. *TSS* denotes the transcription start site of the gene (in base-pair genomic coordinates). Superscript symbols next to molecular traits indicate that the corresponding gene–molecular-trait association also reached RWAS significance in additional AD GWAS summary statistics beyond the primary Bellenguez *et al.*^22^ analysis.*^†^* Kunkle et al. 2019^31^;*^§^* Jansen et al. 2021^32^;‡ Wightman et al. 2021^33^.

RWAS associations showed both shared and molecular-trait-specific patterns (Figure 2B). Several genes, including *CR1*, *BIN1*, *INPP5D*, *TREM2*, *PTK2B*, and *CLU*, were significant in multiple tissues and cell types. In contrast, *ZNF428*, *NECTIN2*, and *APOE* showed strong signals in only one or a few molecular traits, indicating molecular-trait-specific regulatory mechanisms. Overall, 69 of the 138 RWAS-significant genes were associated with a single molecular trait (Table S1). Directional patterns also differed across genes. For example, *CLU* consistently exhibited negative effects across all imputable molecular traits, whereas *APH1B* exhibited positive effects across all molecular traits, consistent with prior functional studies showing their opposite roles in AD patho-genesis^34,35^. Other genes displayed mixed patterns across cell types, such as *PICALM*, which had positive effects in Mono and Oli and negative effects in Mic, Ast, Exc, and AC.

Using the 15,423 imputable genes as background, enrichment analysis of the 138 RWAS-significant genes identified 344 Gene Ontology (GO) terms (*Benjamini–Hochberg* FDR *<* 0.05) using the `gprofiler2` v.0.2.4 `R` package^36^ (Figure 3). These terms grouped the associated genes into AD-relevant biological themes, including immune and complement activity, vesicle/endosomal and membrane-associated biology, and amyloid/lipid handling with glial response. Detailed GO and WikiPathways terms, contributing genes, and interpretation of CNV-annotated WikiPathways results are provided in Supplementary Note S4 and Figure 3; these enrichments should be interpreted as biological context for the associated gene set, rather than direct mechanistic evidence for every contributing gene.

**Figure 3.**
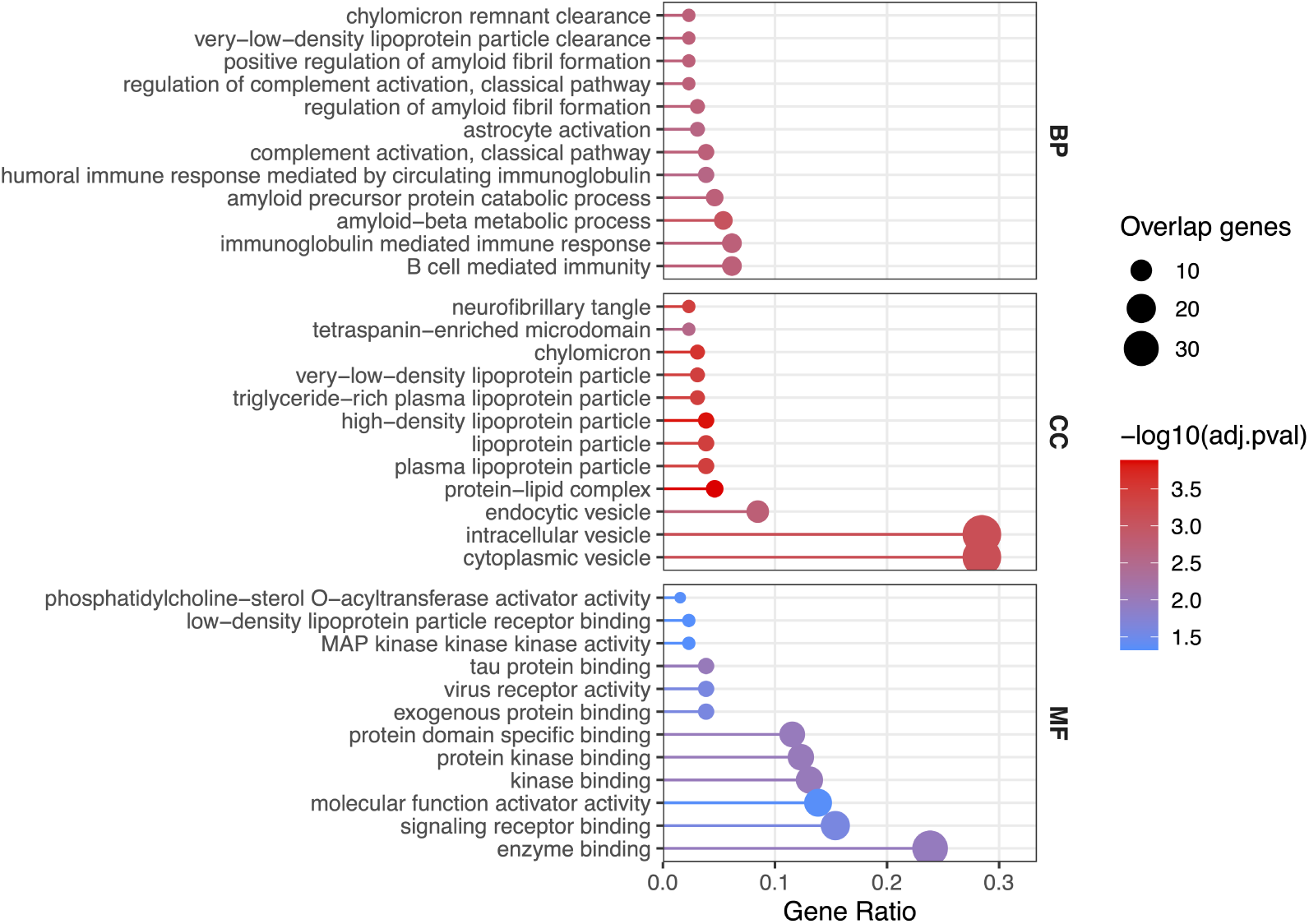
Gene Ontology (GO) Enrichment Analysis of RWAS-Significant Genes. GO enrichment was performed with g:Profiler (organism: hsapiens; sources: Gene Ontology Biological Process (GO:BP), Cellular Component (GO:CC), and Molecular Function (GO:MF)). *p*-values from a hypergeometric test were adjusted by *Benjamini–Hochberg* FDR; the color scale shows − log_10_(FDR). Gene ratio is intersection_size/query_size (*N*query = 138). Point size indicates the number of overlapping genes. The background gene set was restricted to imputable genes (*N*_background_ = 15,423). The plot shows the top 12 terms per ontology by FDR and then gene ratio, highlighting immune and complement activity, amyloid and lipid handling, glial activation, vesicle and endosomal compartments, and receptor or kinase binding terms. Among WikiPathways CNV-associated gene sets, the 16p11.2 proximal deletion set overlapped *YPEL3*, *MAPK3*, *DOC2A*, *TAOK2*, *TMEM219*, *INO80E*, *KCTD13*, *KIF22*, *PPP4C*, *SEZ6L2*, and *ASPHD1*; the 10q22–q23 copy number variation set overlapped *TSPAN14*, *MAT1A*, *DYDC1*, *ADAM10*, and *RSPH6A*.

### Multi-Context Modeling Expands and Refines Alzheimer’s Disease RWAS Signals

Prediction model choice also affected the number and distribution of significant associations (Figure 4). In OPC and Mono, approximately 20–30% of significant RWAS associations were identified only by multivariate models and would have been missed by univariate approaches (Figure 4A), consistent with limited power from the small Mono sample size (*N* = 226) and low OPC cell-type proportion (3%). Specifically, the *PICALM* locus illustrates this dependence on the method, with AC and Exc associations detected exclusively by multivariate methods, and Mic and Ast associations significant across all eight methods (Figure 4B). Across molecular traits, Mic showed the highest significant association proportion (1.20% vs 0.41-0.80% for other traits), defined as the number of RWAS-significant genes divided by the total number of imputable genes in that molecular trait. This was observed despite Mic having the fewest imputable gene–molecular-trait pairs, reinforcing the importance of Mic in AD pathogenesis. Bulk tissue molecular traits such as PCC (0.70%), AC (0.80%), and DLPFC (0.74% for gene expression, 0.71% for protein abundance) overall yielded higher proportions than cell-type contexts (other than Mic), which ranged from 0.41% in OPC to 0.65% in Ast. AC produced the largest number of significant RWAS genes (87), while OPC yielded the fewest (15) (Figure 4C).

**Figure 4.**
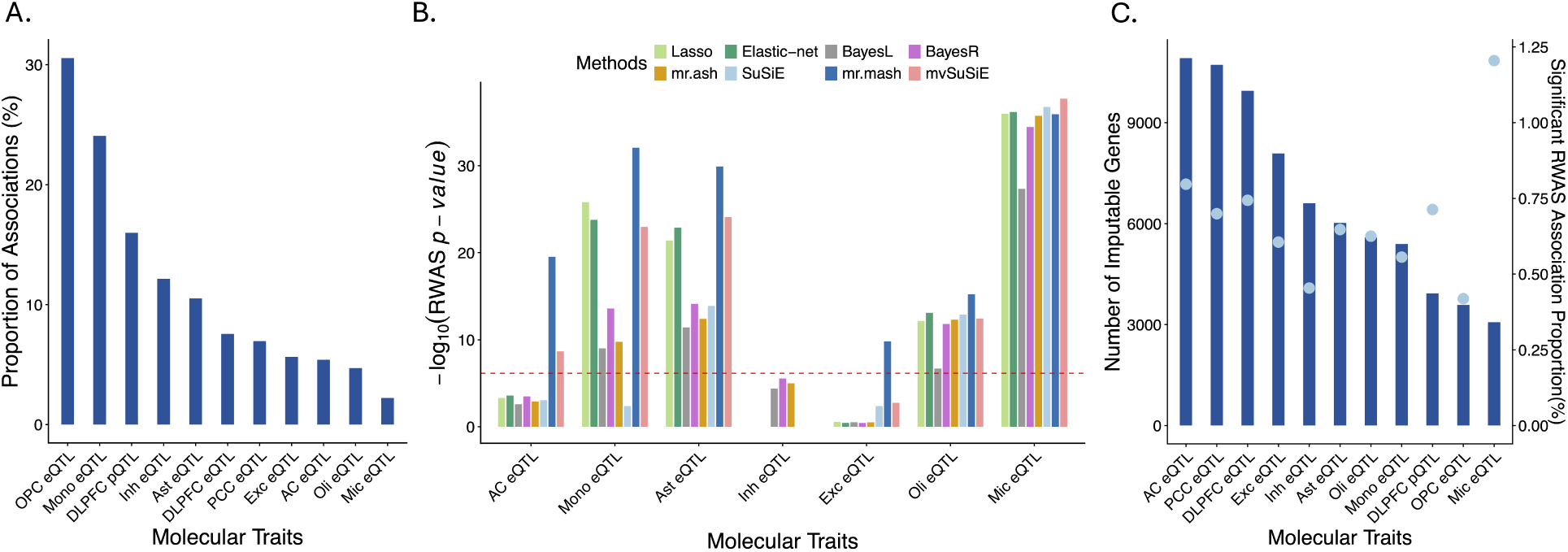
Model Choice and Significant RWAS Associations. (A) Proportion of significant RWAS associations exclusively identified by multivariate methods. (B) Molecular-trait- and method-specific RWAS associations for the *PICALM* gene. (C) Blue bars (left Y-axis) show the number of imputable genes per molecular trait, whereas red points (right Y-axis) show the proportion of significant RWAS associations among imputable gene–molecular-trait pairs for each molecular trait.

The broader RWAS association set still required post-RWAS fine-mapping before they can be interpreted as candidate regulatory mechanisms. It is known that RWAS can produce spurious associations through LD hitchhiking and co-regulation^18^. To refine RWAS associations and prioritize gene–molecular-trait pairs with suggestive evidence of non-zero effects, we applied single-group *cTWAS*^19^ and multi-group *cTWAS* (*M-cTWAS*)^21^. We focused primarily on *M-cTWAS*, which performs joint variant–gene fine-mapping across molecular traits, and used single-group *cTWAS* as a supporting analysis within each molecular trait. Among the 327 RWAS-significant gene–molecular-trait pairs, 86 were prioritized by *M-cTWAS* using a relaxed prioritization criterion, defined as inclusion in a credible set (CS) or PIP *≥* 0.5. These prioritized gene–molecular-trait pairs were interpreted as having suggestive evidence of non-zero effect, rather than definitive causality. Among the total of 86 *M-cTWAS*-prioritized RWAS-significant gene–molecular-trait pairs (corresponding to 43 unique genes), 84 were also supported by single-group *cTWAS* results under the same criteria (Supplementary Table S4). *M-cTWAS* fine-mapping identified 72 CSs overall, with detailed posterior-summary definitions, CS composition, and single-group *cTWAS* comparison provided in Supplementary Note S3 and Supplementary Table S4. Supplementary Figure S8 shows *M-cTWAS* posterior inclusion probabilities (PIP) and CS inclusion status across gene–molecular-trait pairs for genes that were RWAS-significant in at least one molecular trait. *M-cTWAS* therefore refined the broader marginal RWAS set into a more selective set of prioritized signals by jointly modeling variants and correlated gene-level molecular traits.

Across the *M-cTWAS*-prioritized gene sets, many of the strongest signals occur in gene expression from immune and glial contexts (Figure 5), consistent with the well-established role of microglia and immune pathways in AD genetic risk^22^. High-PIP examples include *PICALM* (Mic eQTL), *CR1* (AC eQTL), *RABEP1* (Mic eQTL), *USP6NL* (Mic eQTL), and *CASS4* (Mic eQTL). We also observed multiple high-PIP genes linked to amyloid processing or endolysosomal trafficking, such as *APP* (Ast eQTL)^37^, *CR1* (AC eQTL)^38^, *PICALM* (Mic eQTL)^39^, and *RABEP1* (Mic eQTL)^40^. Several other prioritized genes, including *TP53INP1* (Mic eQTL)^41^, *RHOH* (Mic eQTL)^42^, and *WDR81* (DLPFC eQTL)^43^, may point to pathways related to cellular stress response, immune signaling, and endolysosomal function, respectively, although their roles in AD are less firmly established. Together, these patterns indicate that the subset of RWAS associations retained after fine-mapping is enriched for biologically coherent immune, glial, and vesicle-related pathways.

**Figure 5.**
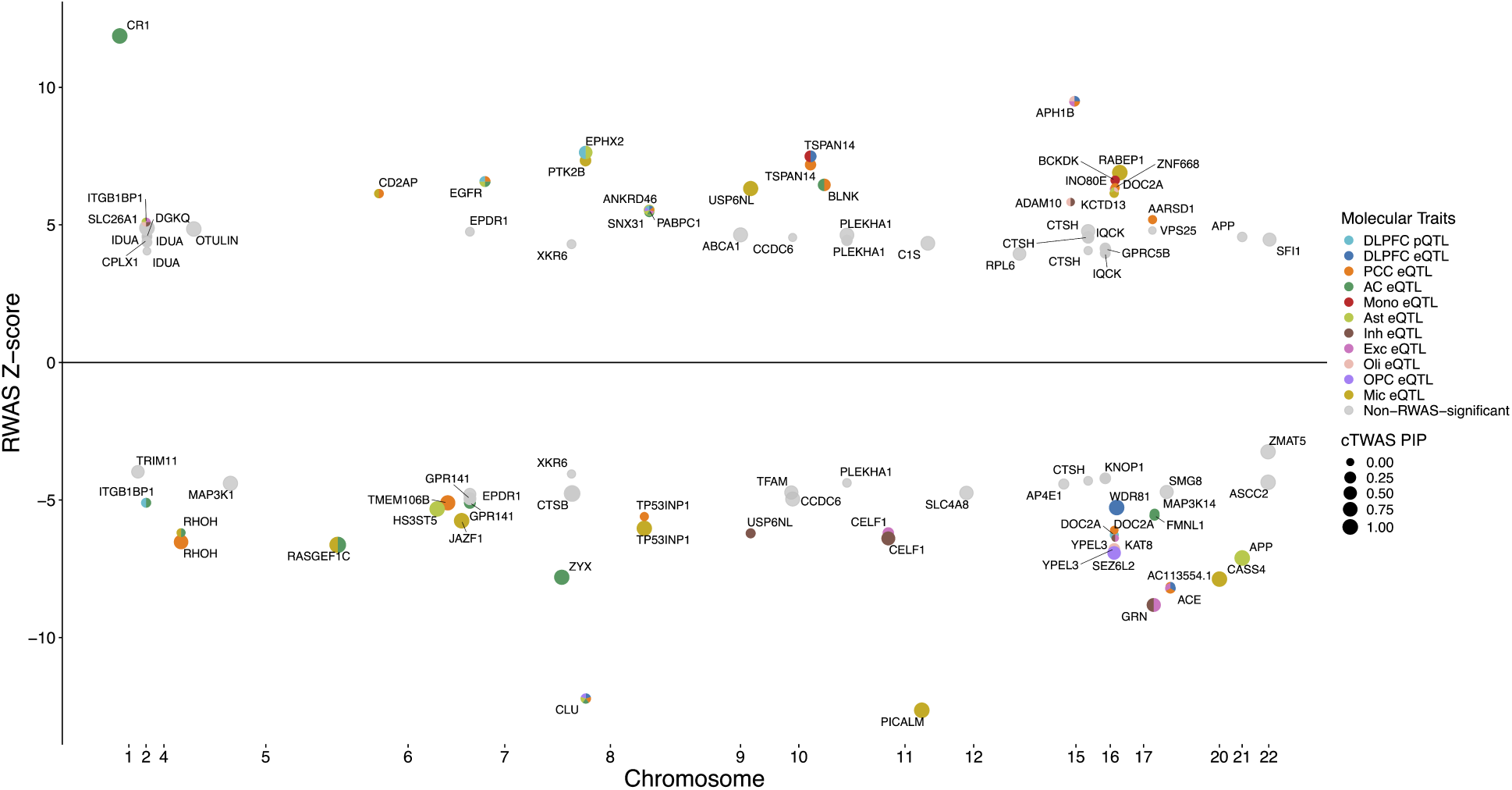
*M-cTWAS* Variant–Gene Fine-Mapping of RWAS Signals. RWAS z-scores are shown for gene–molecular-trait pairs that were prioritized by *M-cTWAS* PIP ≥ 0.5 or inclusion in a credible set. Point color indicates molecular traits. Point size encodes the PIP (range 0–1; larger points indicate higher PIP). Grey points denote gene–molecular-trait pairs that do not surpass the RWAS Bonferroni threshold (*p <* 0.05/73,874), whereas colored points are RWAS-significant. *M-cTWAS* fine-mapping refines RWAS associations to a subset of gene–molecular-trait pairs with stronger fine-mapping support, reducing spurious signals driven by LD or co-regulation. A corresponding RWAS z-score plot for gene–molecular-trait pairs prioritized by single-group *cTWAS*results is shown in Supplementary Figure S6.

### Locus-Level Fine-Mapping Separates Regulatory Signals from LD Hitchhiking

We used representative loci to show how fine-mapping after RWAS can detect LD hitchhiking, prioritize molecular context, resolve correlated nearby genes, and flag loci that require caution because of LD mismatch (Figure 6). At the *TREM2* locus (Figure 6A), marginal RWAS identified *TREM2* associations in several molecular traits, but *M-cTWAS* shifted support away from the gene and assigned high PIP to the nearby GWAS variant rs187370608. This pattern is consistent with LD hitchhiking^44^ and does not argue against *TREM2* biology, but suggests that the marginal RWAS signal is more consistent with local variant effects than with baseline *cis*-regulated *TREM2* expression in the contexts tested. This interpretation is biologically plausible because rare *TREM2* coding variants influence AD risk, and *TREM2* biology affects microglial lipid and apolipoprotein/A*β* handling, plaque-associated microglial states, metabolic fitness, and inflammatory signaling^45–50^. Consistent with this interpretation, *TREM2* was not identified as a DLPFC *cis*-eGene in GTEx or ROSMAP^3,51^, our *cis*-eQTL analysis across AC, DLPFC, Mic, and PCC tested approximately 13,000 variants in the *TREM2* locus and found no significant *cis*-eQTL after FDR correction, and *ColocBoost* showed no shared variants between *TREM2* expression and AD risk^30^. The *M-cTWAS*-prioritized variant rs187370608 is also a previously established AD risk variant^52^. Together, these observations support LD hitchhiking rather than baseline-expression mediation at this locus.

**Figure 6.**
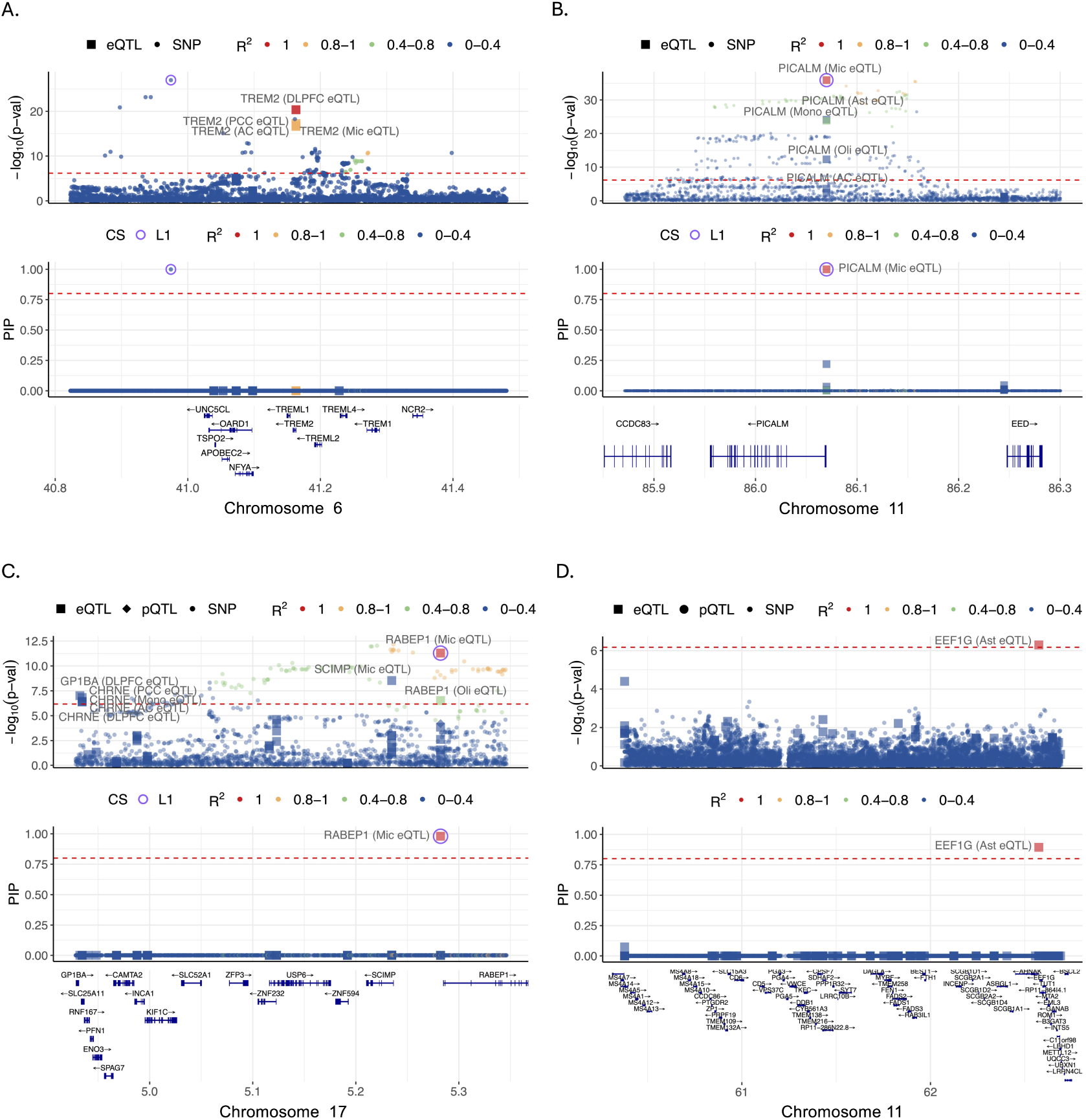
Representative *M-cTWAS* Locus Plots. The locus plot compares RWAS and *M-cTWAS* association results at a locus across multiple molecular traits. The upper panel shows RWAS − log_10_(*p*) for genes (squares) and GWAS − log_10_(*p*) for variants (small dots), with each modality type represented by a different shape. The lower panel shows *M-cTWAS* posterior inclusion probabilities (PIPs) for the same set of SNPs and genes. In both panels, point colors indicate the correlation *r*^2^ between each gene or SNP and the focal gene (the gene with the highest PIP in the locus), computed from the regional SNP-gene and gene-gene correlation matrices. Colored circles/rings indicate credible set (CS) inclusion status, with different circle colors denoting different CSs, whereas SNPs or genes without a circle are not assigned to any CS. These plots illustrate the local LD/co-regulation structure surrounding the focal gene and prioritization of candidate gene–molecular-trait pairs and SNP signals. (A) At the *TREM2* locus, marginal RWAS associations suggest multiple significant genes; however, *M-cTWAS* fine-mapping assigns high posterior inclusion probability (PIP) to a nearby SNP, and the gene signal disappears. (B) At the *PICALM* locus, the Mic eQTL signal is supported by both RWAS association and *M-cTWAS*, whereas Ast and Mono signals drop after fine-mapping. (C) At the *RABEP1* locus, marginal RWAS identified multiple significant associations across molecular traits, including *RABEP1* –Mic eQTL, *SCIMP* –Mic eQTL, *GP1BA*–DLPFC eQTL, and *CHRNE*. However, after multi-context joint variant–gene fine-mapping with *M-cTWAS*, only *RABEP1* –Mic remained prioritized (PIP ≈ 0.98) while being included in a CS. This suggests that although several nearby gene–molecular-trait pairs show marginal association, the *RABEP1* –Mic eQTL signal best explains the locus after accounting for LD and co-regulation across molecular traits. (D) At the *EEF1G* locus, *M-cTWAS* prioritizes *EEF1G*–Ast as the top fine-mapped signal (PIP ≈ 1), whereas *MS4A7* –Mic shows a substantially lower PIP (≈ 0.1). Because the GWAS association pattern failed the LD mismatch diagnostic against the reference LD structure, this locus was fine-mapped without LD, and CS information is not displayed because it is not directly comparable to the LD-based CSs reported for other loci.

At the *PICALM* locus (Figure 6B), fine-mapping distinguished the molecular context most strongly supported by the data. Predicted *PICALM* expression was RWAS-significant in Ast, Mic, Mono, and Oli, but Mic eQTL was the only molecular trait with high *M-cTWAS* PIP. This prioritization is supported by *ColocBoost*, where *PICALM* Mic eQTLs and AD GWAS signals were included in the same 95% Colocalization Set (CoS), a cluster of signals grouped as sharing colocalized genetic association evidence^30^. The Mic eQTL interpretation was further supported by replication across three additional AD GWAS cohorts^31–33^ and recent functional evidence showing that a *PICALM* AD risk allele reduces PU.1 binding and *PICALM* expression in iPSC-derived microglia, impairing A*β* and myelin-debris uptake and promoting lipid-droplet accumulation^53^. Although Ast and Mono eQTLs also joined 95% CoSs with Mic, the combined RWAS, colocalization, and *M-cTWAS* evidence most strongly implicated Mic eQTL as the primary regulatory molecular trait among the 11 molecular traits tested. This illustrates the added value of *M-cTWAS* over colocalization alone for prioritizing among correlated molecular-trait predictors.

At the *RABEP1* locus (Figure 6C), multiple nearby genes and molecular traits showed marginal RWAS associations. *M-cTWAS* resolved these correlated signals by prioritizing *RABEP1* –Mic eQTL, while other *RABEP1* and *SCIMP* molecular traits were assigned near-zero PIPs. This prioritization is supported by *ColocBoost*, where the *RABEP1* Mic eQTL and AD GWAS signal were included in the same 95% CoS^30^. It is also consistent with the established role of *RABEP1* (Rabaptin-5) as a Rab5 effector in early endosome fusion^54^ and with more recent evidence placing Rabaptin-5 in early-endosome homeostasis and damaged-endosome autophagy^55^, together with rare-variant evidence in AD linking *RABEP1* p.R845W to enlarged early endosomes, endocytosis dysregulation, and increased toxic amyloid-*β* accumulation^40^. This rare coding variant provides mechanistic support for *RABEP1* biology, although it should not be interpreted as direct evidence for the regulatory RWAS allele. By contrast, *SCIMP* is a plausible nearby immune gene, but is primarily characterized as an adaptor in immune receptor signaling^56^. Thus, this locus illustrates how cross-context fine-mapping can concentrate support on one gene–molecular-trait pair within a broader correlated regulatory signal.

At the *EEF1G* locus (Figure 6D), fine-mapping prioritized a previously unreported *EEF1G* –Ast association over *MS4A7* –Mic, even though the latter is known AD-associated gene. The *EEF1G* –Ast signal had much higher PIP (*≈* 1) than *MS4A7* –Mic (PIP *≈* 0.1), and the predicted expression correlation between the two signals was low (*r*^2^ *≈* 0.0045), suggesting that they may tag different underlying association signals. However, this region failed the LD mismatch diagnostic and was fine-mapped without LD under a single-effect setting (*L* = 1). Direct mechanistic evidence for *EEF1G* in AD remains limited; *EEF1G* encodes a component of the eukaryotic elongation factor 1 complex, and broader elongation-factor literature implicates impaired de novo protein synthesis in AD-relevant synaptic biology^57^. The *EEF1G* –Ast association should therefore be interpreted as hypothesis-generating, especially because residual shared-variant effects between *MS4A7* and *EEF1G* cannot be ruled out.

## DISCUSSION

FunGen-xQTL Multi-Brain (FGMB) provides a multi-omics regulome-wide association atlas, an eight-method prediction framework, and a joint variant–gene fine-mapping layer for aging brain genetic analyses. FGMB is designed to connect GWAS signals not only to genes, but also to the molecular modality, brain region, cell type, and cell subtype in which those signals are most strongly supported. Although the AD analysis is the main application here, the atlas itself is designed for broader studies of neurodegenerative and aging-related brain traits, with reusable prediction weights and joint variant–gene fine-mapping across molecular contexts. This structure allows disease-associated regulatory hypotheses to be evaluated at the level of specific molecular traits and cellular contexts, rather than being collapsed into a single bulk-tissue signal.

Recent cell-type-aware RWAS studies have been performed using ROSMAP DLPFC single-nucleus molecular measurements and AD GWAS summary statistics^58,59^. These study-specific analyses reinforce the value of resolving genetic regulation at cell-type and cell-subtype resolution. FGMB was developed independently for a different scale and purpose as a FunGen-AD atlas that releases reusable prediction weights, RWAS association results, and joint variant–gene fine-mapping outputs across 36 molecular datasets, 18 contexts, and 3 modalities. Compared with these single-nucleus RWAS analyses, FGMB uses larger ROSMAP-derived single-nucleus references, including a CUIMC1/CUIMC2/MIT mega-analysis for six major cell types (*N* = 733–737), integrates bulk and single-nucleus brain, peripheral monocyte, splicing, and protein datasets from ROSMAP, MSBB, and Knight-ADRC, and applies a broader statistical framework with eight prediction methods and single- and multi-context joint variant–gene fine-mapping. This combination gives the atlas greater molecular coverage, stronger prediction performance, and more selective prioritization of AD-associated gene–molecular-trait signals.

FGMB is also a protocol and software resource. The `xqtl-protocol` workflow and the `pecotmr R` package provide a reproducible path from harmonized genotype and molecular data to prediction weights, RWAS association statistics, and both single-group *cTWAS* and multi-group *M-cTWAS* joint variant–gene fine-mapping outputs (see Code Availability). The predictor-building component supports eight sparse-regression, Bayesian, and multivariate models and selects weights by cross-validation within each molecular dataset. In the primary AD application, expanding beyond *elastic-net* and *lasso* prediction methods added 28,014 imputable gene–molecular-trait pairs and 7,327 unique genes, increased selected-weight median cross-validation *R*^2^ from 0.0203 to 0.0438, and reached median *R*^2^ = 0.0455 when selected among multivariate Bayesian methods (*mr.mash* and *mvSuSiE*). These improvements show that model selection is a practical route to producing more accurate RWAS weights when reference panels are modest in size and regulatory effects may be shared across contexts.

The AD application illustrates why this combination of molecular coverage and model selection matters. Using 11 ROSMAP molecular datasets with Bellenguez *et al.* AD GWAS summary statistics, FGMB identified 327 RWAS-significant gene–molecular-trait pairs corresponding to 138 unique genes. Of these genes, 42 had not been previously implicated by GWAS, RWAS, or curated AD gene resources (Table 2). Their pathway enrichments pointed to processes already central to AD biology, including immune and complement activity, endosomal and vesicle trafficking, membrane localization, and cell-surface organization. These RWAS findings should be interpreted as molecular-trait-specific association hypotheses, while highlighting that the atlas can recover biologically coherent signals beyond previously catalogued AD genes.

The joint variant–gene fine-mapping analysis adds a second layer of resolution by asking which RWAS associations remain supported after modeling nearby variants, genes, and correlated molecular traits together across contexts. *M-cTWAS* prioritized 86 of the 327 RWAS-significant gene–molecular-trait pairs, corresponding to 43 unique genes, with global prior estimates suggesting stronger enrichment for microglial and selected bulk brain molecular traits than for several other contexts (Figure S7). Many credible sets were dominated by SNP signal rather than gene signal. This pattern is consistent with recent literature showing that some marginal RWAS associations may reflect LD hitchhiking, co-regulation, cross-context tagging, or regulatory mechanisms not captured by the measured molecular traits, rather than a supported gene-level effect. In this way, *M-cTWAS* is not simply a follow-up filter for significant RWAS hits; it is part of the resource rationale, allowing users to separate broad molecular-trait association discovery from more conservative variant–gene prioritization.

Several considerations should guide the use of FGMB. First, the TAD-boundary-enhanced *cis* window was intentionally extended to capture broader *cis*-regulatory architecture^60,61^. This design can improve prediction, but some high-performing models are non-sparse, leaving their downstream use sensitive to variant coverage in the target GWAS summary statistics and exposing RWAS z-scores to LD tagging by non-trait-relevant variants^20^. Users can mitigate this issue by harmonizing and imputing GWAS summary statistics to the provided LD reference panel, or by using released weights from more sparse models. We note that *mvSuSiE*, the highest-performing prediction method in our analyses, is a sparse Bayesian fine-mapping model and may therefore be more robust than diffuse non-sparse predictors to both variant-coverage gaps and LD tagging. Second, FGMB includes brain regions, major brain cell types, selected cell subtypes, monocytes, and three regulatory modalities, but finer cell-state and cell-subtype resolution remains limited by available sample size, particularly for microglial and astrocytic subtypes. The *M-cTWAS* results themselves point to this remaining gap. If the relevant molecular contexts were fully captured, more posterior support would be expected to concentrate on gene–molecular-trait predictors; instead, many credible sets in our analysis were still dominated by SNP signal. This pattern may reflect residual LD hitchhiking, cross-context tagging, regulatory mechanisms acting through molecular traits or cell states not yet represented in FGMB, or non-regulatory variant effects. This observation motivates the next stage of subtype-level and cross-population single-cell reference data generation by FunGen-AD. Third, while *cTWAS* and *M-cTWAS* provide more conservative prioritization, they are known to lack power under some settings^19,21^. The prioritized gene–molecular-trait pairs should therefore be viewed as signals with fine-mapping support, not as definitive causal assignments. Fourth, interpretation remains difficult in genomic regions with exceptionally strong or complex association and LD patterns. For example, the *APOE* region has a dominant AD association signal and can be sensitive to the LD reference panel used for RWAS and fine-mapping; the *MAPT* region contains a complex extended haplotype structure; and the 16p11.2 breakpoint 4–breakpoint 5 (BP4–BP5) copy-number-variant region contains multiple nearby genes that can be difficult to separate using association data alone. We used a large ADSP/GCAD-based LD reference panel and LD mismatch diagnostics, but locus-level signals in these regions should still be interpreted with caution. Finally, the current reference panels from ROSMAP, MSBB, and Knight-ADRC were derived predominantly from individuals of European ancestry. Potential ancestry mismatch will affect association testing, thus use in non-European GWAS cohorts may be limited by differences in LD structure, allele frequencies, and regulatory architecture. FunGen-AD is generating new multi-ancestry functional-genomic reference data, and future FGMB releases are designed to incorporate these data as they become available.

In summary, FGMB provides reusable RWAS atlas and scalable analysis framework for aging brain genetic analyses. It links GWAS signals to molecular traits, contexts, and candidate genes while separating marginal RWAS associations from variant-driven signals. The same resource framework has already supported companion applications to aging and telomere length, and to neuroimaging-derived endophenotypes^62,63^. However, one key contribution of this work is methodological because the developed workflow can be rerun as new molecular datasets become available, extended to additional modalities or ancestries, and adopted by other groups to build comparable RWAS resources. By making model training, model selection, association testing, and joint variant–gene fine-mapping part of one reproducible pipeline, FGMB provides a practical route from functional-genomic data to regulome-wide association resources that can be updated, audited, and tested experimentally.

## METHODS

Throughout the Methods, we use molecular dataset to refer to a source-specific molecular measurement panel used for predictor training, such as DLPFC eQTL, CUIMC1 Ast eQTL, or MIT Mic.12 eQTL. We use context for the tissue, brain region, cell type, or cell subtype; regulatory modality for the molecular phenotype being modeled (*e.g.*, gene expression, protein abundance, or splicing); and molecular trait for the phenotype modeled within a gene-level association or fine-mapping analysis. Upstream cohort organization, genotype processing, molecular phenotype preprocessing, xQTL association testing, TAD-boundary-enhanced xQTL windows, and harmonized release formats are described in the companion FunGen-xQTL Atlas descriptor^64^. Here, we start from those harmonized molecular datasets to train FGMB prediction models, evaluate model availability and prediction performance, perform AD RWAS, and carry out joint variant–gene fine-mapping.

### Multi-Omics Reference Data Processing

We used genotype and molecular data that were preprocessed, harmonized, and curated by FunGen-AD for well-characterized source datasets, including ROSMAP^23^, MSBB^24^, and Knight-ADRC (Charles F. and Joanne Knight Alzheimer’s Disease Research Center)^25^. FGMB used harmonized molecular datasets spanning gene expression, protein abundance, and splicing across bulk brain regions, peripheral monocytes, single-nucleus cell types, and single-nucleus cell subtypes. The ROSMAP-derived datasets included bulk RNA-seq from DLPFC, PCC, and AC; DLPFC proteomics; bulk splicing models in DLPFC, PCC, and AC; monocyte expression; CUIMC1; the MIT single-nucleus dataset described by Xiong *et al.* 2023^29^; and the CUIMC1/CUIMC2/MIT mega-analysis, where CUIMC2 contributed recently generated ROSMAP DLPFC single-nucleus multiome RNA profiles^5,26–28^. Additional molecular datasets came from MSBB bulk RNA-seq and proteomics and from Knight-ADRC parietal cortex RNA-seq and proteomics. Dataset-level composition, sample sizes, source-study details, and extended resource summaries are provided in Supplementary Note S1, Table 1, and the companion FunGen-xQTL Atlas descriptor^64^. For splicing predictors, we used LeafCutter2-derived productive (PR) and unproductive (UP) splicing traits and excluded events flagged as non-informative or exclusionary, such as inconclusive (“IN”) or not expressed (“NE”)^28^.

### Genotype Data Processing

Source-level genotype processing and QC for the molecular reference datasets are described in the companion FunGen-xQTL Atlas descriptor^64^. For FGMB predictor training, we applied sample-aligned genotype filters to ROSMAP, MSBB, and Knight-ADRC based on minor allele frequency (MAF *>* 0.0025), Hardy–Weinberg equilibrium (HWE; *p >* 1 *×* 10^−8^), minor allele count (MAC *>* 5), variant missingness rate (imiss *<* 1.0)^65^, and a minimum per-variant genotype variance threshold (x_variance *>* 0.01). Per-variant genotype variance metrics from this step were carried forward to the RWAS predictor-training stage and used for subsequent variance-based variant filtering. To preserve interpretability after residualization and optional scaling, we generated separate scaling factors for the genotype and molecular-trait matrices (X_scalar and Y_scalar) to rescale model coefficients to their original units. Samples with *>* 10% missing genotype data across all variants were removed, and only samples with complete data across covariates, molecular profiles, and genotype data were retained.

### Training Molecular Trait Predictors for Multi-Context, Multi-Omics Data

Prediction models were trained within the TAD-boundary-enhanced *cis* windows defined by the companion FunGen-xQTL Atlas workflow^64^. In brief, the workflow merges human hippocampus and cortex TAD maps^66^, constructs generalized TAD-boundary domains, combines relevant TAD-boundary domains with the standard *±*1 Mb gene-centered *cis* window, and winsorizes extremely large regions at the 99th percentile of genome-wide region length, corresponding to approximately 20 Mb. FGMB analyses were restricted to protein-coding genes.

We trained molecular trait predictors (*i.e.*, RWAS weights) using genotype and molecular trait data from each molecular reference panel. The RWAS weight training workflow was implemented via `xqtl-protocol` together with the `pecotmr` v.0.3.4 `R` package. For each gene–molecular-trait pair, we fitted different models to estimate predictors that capture genetic regulatory effects. We employed a total of eight modeling approaches, including six univariate methods that model one molecular trait at a time, and two multivariate methods that model multiple molecular traits jointly. The univariate methods were:

- *SuSiE* (`susieR`, v.0.14.7)^67^. This is a Bayesian variable selection method developed for fine-mapping that models regression coefficients as a sum of sparse single effect vectors. This method is suitable for very sparse genetic architectures. The initial number of single effects to consider (*L*) was set to 5, and *L* was increased by 5 each time the limit was reached, with a maximum *L* of 20.
- *mr.ash* (`mr.ash.alpha`, v.0.1-43)^14^. This is a Bayesian approach that uses adaptive shrinkage priors with a flexible mixture of normal distributions on the regression coefficients and applies variational approximations to improve predictive accuracy and computational efficiency. This method is suitable for sparse and moderately dense genetic architectures. We used data-driven standard deviations to compute the prior variances (`sa2`) for the mixture components.
- *lasso* (`glmnet`, v.4.1-8)^68,69^. This is a regularized regression method that imposes an L1 penalty on the regression coefficients to select a sparse subset of predictors by shrinking the remaining regression coefficients exactly to zero. This method is suitable for sparse and moderately dense genetic architectures.
- *elastic-net*^69,70^. We implemented the *elastic-net* model with `glmnet` v.4.1-8 and mixing parameter *α* = 0.5, combining *L*_1_ and *L*_2_ penalties to handle correlated predictors such as variants in LD. This method is suitable for sparse and dense genetic architectures and is used in the original *PrediXcan* implementation^6^.
- *BayesL* (`qgg`, v.1.1.6)^71,72^. This is a Bayesian approach that uses a double exponential prior on the regression coefficients. This method is suitable for dense genetic architectures, as the prior does not perform variable selection. The total number of iterations was set to 5,000 with a burn-in of 1,000 and a thinning interval of 5.
- *BayesR* (`qgg`, v.1.1.6)^72,73^. This is a Bayesian approach that models the regression coefficients using a four-class mixture of normal distribution priors, allowing for a mixture of small, moderate, and large effects. This method is suitable for sparse and dense genetic architectures. As with *BayesL*, we set the total number of iterations to 5,000 with a burn-in of 1,000 and a thinning interval of 5 for *BayesR*. The two multivariate methods were:
- *mr.mash* (`mr.mash.alpha`, v.0.3.36)^15^. This is the multivariate version of *mr.ash*, and it employs a flexible mixture of multivariate normal priors to leverage the sharing of genetic effects across molecular traits. We used the mixture component weight cutoff (`w0_threshold`) as 1 *×* 10^−8^ and enabled residual covariance updates (`update_V = TRUE`), with convergence tolerance (*tol*) of 0.01.
- *mvSuSiE* (`mvsusieR`, v.0.1.7)^74^. This is the multivariate version of *SuSiE*, and it is a sparse Bayesian fine-mapping model that employs a flexible mixture of multivariate normal priors to leverage the sharing of genetic effects across molecular traits. The maximum number of non-zero effects (*L*) was set to 30.

At the predictor-training stage, genotype quality-control thresholds were reapplied to the genotype matrices after restricting to samples with complete molecular trait and covariate data. In addition to the standard genotype QC filters, we applied minor allele frequency cutoff (`min_twas_maf` = 0.01) and a per-variant genotype variance cutoff (min_twas_xvar = 0.01).

Coefficient scaling also differed between the two workflows. In the univariate workflow, prediction models were fitted using covariate-residualized molecular-trait values together with the corresponding sample-aligned genotype matrix, and coefficient rescaling was handled separately through `X_scalar` and `Y_scalar`. In the multivariate workflow, the shared genotype matrix was modeled jointly with the residualized multi-molecular-trait matrix using scaling factors fixed at 1; therefore, no additional coefficient rescaling was applied, and coefficients were retained on the scale of the fitted multivariate model.

The two multivariate methods require user-specified estimates of the covariance matrices of genetic effects across molecular traits. To compute these mixture priors, we adopted the *Extreme Deconvolution* framework developed by Bovy *et al.*^75,76^, which computes both data-driven and canonical covariance matrices. *mvSuSiE* also requires estimates of the weight associated with each prior covariance matrix. To obtain these estimates, we fitted *mr.mash* — which estimates the weights during model fitting — with the covariance matrices from the previous step. After removing covariance matrices with very small estimated weights (*w*_0_ *≤* 1 *×* 10^−4^), the remaining covariance matrices and associated weights were passed to *mvSuSiE*^74^ as the mixture prior. The residual covariance estimated from *mr.mash* was used in *mvSuSiE*. A representative visualization of the retained mixture prior components and their estimated weights for the ROSMAP 11-context analysis presented in the main text is shown in Figure S9.

We used five-fold cross-validation to evaluate model performance across the eight methods. Performance was based on *R*^2^ and the *p*-value from the linear regression of the observed molecular trait values on the predicted molecular trait values. A gene–molecular-trait pair was considered imputable if it achieved a cross-validation *R*^2^ *≥* 0.01 and a corresponding *p <* 0.05 from at least one of the eight methods^16^. Gene–molecular-trait pairs that did not meet this threshold were excluded from downstream analyses to ensure the reliability of imputed molecular trait estimates. The molecular trait prediction models were trained on the full dataset to generate final predictors, while cross-validation was used solely for model performance comparison and determining imputability. We used cross-validation to select among prediction methods. Downstream RWAS testing used the established summary-based statistic with prediction weights, GWAS z-scores, and LD matrices harmonized to the same variant set and allele orientation. For fine-mapping, the LD reference panel and the `cTWAS` LD mismatch diagnostic were used to flag loci where long-range or non-sparse predictors could be more vulnerable to LD mismatch or tagging.

### LD Reference Data and GWAS Summary Statistics

Our LD reference data were derived from whole-genome sequencing data of 16,571 non-Hispanic white individuals provided by the Genome Center for Alzheimer’s Disease (GCAD)^77^. LD matrices were computed for 1,361 approximately independent blocks, using boundaries derived from the 1000 Genomes Project Phase 3 European (EUR) ancestry reference panel^78^. Before LD computation, the genotype data were filtered to retain variants with MAF *>* 0.05%, MAC *>* 5, and missing rate *<* 5%^77^.

We used summary statistics from several Alzheimer’s disease GWAS, including the large-scale meta-analysis by Bellenguez *et al.*^22^ (111,326 cases and 677,663 controls), Jansen *et al.*^32^ (71,880 cases and 383,378 controls), Kunkle *et al.* stage 1^31^ (21,982 cases and 41,944 controls), and Wightman *et al.*^33^ (90,338 cases and 1,036,225 controls). To facilitate cohort-specific validation, we incorporated five additional sub-cohort GWAS datasets. Three came from subsets of the Bellenguez *et al.* study: the European Alzheimer & Dementia Biobank (EADB)^79^, which aggregates data from 15 European countries; the European Alzheimer’s Disease Initiative (EADI)^80^; and the GR@ACE study (Genome Research at Fundacio ACE)^81^, a Spanish cohort focused on the genetic and environmental risk of Alzheimer’s disease. The remaining two were versions of the Wightman *et al.* dataset, one excluding both UK Biobank and 23andMe samples and one excluding only 23andMe data.

The SNP identifiers were lifted over to the same genome build (GRCh38) as our genotype data. To ensure consistency across data sources, GWAS summary statistics were harmonized to the LD reference panel, aligning variant effect directions relative to the same reference allele across LD, GWAS, and molecular datasets. We removed SNPs that were non-overlapping between summary statistics and LD, strand-ambiguous (e.g., A/T or C/G alleles without sufficient allele frequency context), or duplicated within the merged dataset.

### RWAS Association Testing

We then tested the association of imputed molecular trait values with AD risk using the same test statistic as the summary-based FUSION TWAS framework^7^. For each gene–molecular-trait pair, the RWAS z-scores were computed by integrating variant-level GWAS z-scores (**z**), molecular-trait-specific prediction weights (**w**), and the local LD matrix (**R**) among SNPs.

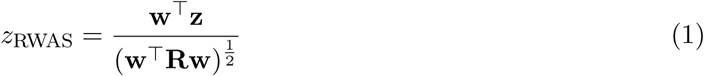

We performed this analysis for all eight methods at each imputable gene–molecular-trait pair and for each molecular dataset from ROSMAP, MSBB, and Knight-ADRC. A Bonferroni-corrected *p*-value threshold (0.05*/M*) was applied to each analysis to define statistically significant RWAS associations, where *M* is the number of imputable gene–molecular-trait pairs.

### Functional Enrichment Analysis

To investigate the biological pathways represented by the RWAS-significant genes, we performed functional enrichment analysis using the `gprofiler2`^36^ v.0.2.4 `R` package (organism: *Homo sapiens* (hsapiens)). Using the full set of 15,423 imputable genes in ROSMAP as the custom statistical background, we queried 138 RWAS-significant genes (Bonferroni-adjusted *p <* 6.77*×*10^−7^). Enrichment analyses used the default `gprofiler2` hypergeometric over-representation test. Significance was determined using the *Benjamini–Hochberg* false discovery rate (BH-FDR), and only terms with FDR *<* 0.05 were considered significantly enriched.

### *cTWAS* and *M-cTWAS* Variant–Gene Fine-Mapping

RWAS is susceptible to false positive discoveries when prediction weights are correlated with nearby genes or variants^18^. These challenges are amplified when multiple molecular traits are integrated because of co-regulation and LD. Multi-group causal TWAS (*M-cTWAS*)^21^ aims to mitigate false positives from confounding effects by jointly modeling the effects of all genes and SNPs within a genomic region across multiple molecular traits. To assess variant and gene-level signals underlying our RWAS associations, we performed *M-cTWAS* analysis using the `cTWAS` v.0.4.21 `R` package, implemented through the integrated `xqtl-protocol twas_ctwas` workflow. *M-cTWAS* was conducted on a per-LD-region basis, with each LD region fine-mapped using all SNPs and genes whose transcription start sites (TSSs) were located within the boundaries of each LD block. Although RWAS association testing was performed across all imputable gene–molecular-trait pairs from the ROSMAP, MSBB, and Knight-ADRC resources, *M-cTWAS* fine-mapping was conducted as a focused application using the 11 ROSMAP molecular datasets included in the AD demonstration analysis.

To estimate the global group prior for SNPs and molecular traits, the RWAS z-scores, molecular trait predictors, and GWAS data were merged genome-wide across all LD regions. The RWAS z-scores were passed as gene-level input (`z_gene`) and the harmonized GWAS z-scores as SNP-level input (`z_snp`) into the *M-cTWAS* model for fine-mapping of candidate variant and gene-level signals. For each gene–molecular-trait combination, we used the RWAS z-scores from the prediction method with the highest cross-validation *R*^2^ and *p <* 0.05. RWAS association results were generated across all prediction methods, but *cTWAS* fine-mapping required a single predictor per gene–molecular-trait pair. Thus, model selection was applied at the RWAS weight training stage to identify the strongest available predictor for downstream *cTWAS* analysis. To ensure consistency for the LD-based fine-mapping model, we scaled the molecular trait predictors by the standard deviation of each SNP estimated from the LD reference panel. We used `prior_var_structure = ‘‘shared_all’’` for global prior training, which assumes a common variance parameter shared across groups. We set `thin = 1.0` to include effects from all SNPs for each gene. The estimated global group prior was then incorporated into the *M-cTWAS* fine-mapping analysis for LD regions containing at least one SNP with GWAS *p <* 5 *×* 10^−8^ or with the sum of gene-level posterior inclusion probability (PIP) *>* 0.5 (min_nonSNP_PIP = 0.5). We defined credible sets using a 95% posterior coverage threshold and applied a minimum absolute correlation *>* 0.1 for variables in a credible set to prioritize genes and SNPs with strong fine-mapping support.

After the initial *cTWAS* fine-mapping step, we applied the `cTWAS` built-in LD mismatch diagnosis using GWAS z-scores, LD reference matrix, and the corresponding SNP annotation files. SNPs or genes showing substantial discrepancy between the observed association pattern and the LD structure were flagged as problematic. For regions containing such genes or SNPs with strong fine-mapping support (PIP *≥* 0.5), we replaced the fine-mapping results with results from fine-mapping performed without LD, using the `cTWAS finemap_regions_noLD()` function.

We additionally performed single-group *cTWAS* to complement the *M-cTWAS* analysis and evaluate molecular-trait-specific fine-mapping results. Single-group *cTWAS* estimates effects for a single molecular trait at a time, enabling independent investigation of molecular-trait-specific signals in loci^19^. To ensure comparability with the *M-cTWAS* analysis, we ran single-group *cTWAS* using the same input data and used the same `twas_ctwas` pipeline from `xqtl-protocol`, with `multi group = FALSE` to perform single-group *cTWAS* and `multi_group = TRUE` to perform *M-cTWAS*. We also used the same settings for global prior training and fine-mapping in both analyses, including `prior_var_structure = ‘shared_all’`, `min_nonSNP_PIP = 0.5`, and `thin = 1.0`, so that SNP effects were included for each gene during fine-mapping.

## Data Availability

The complete RWAS prediction models, Alzheimer’s disease RWAS association results, and joint variant–gene fine-mapping results are available through the Synapse portal under the following accessions: RWAS prediction models, syn69670600; Alzheimer’s disease RWAS association results, syn74908416; and single-context and multi-context joint variant–gene fine-mapping results, syn74908389. The linkage disequilibrium (LD) reference panel used for RWAS association testing and fine-mapping was built from whole-genome sequencing data from participants of European ancestry in ADSP Release 4 and is available through Synapse, syn75082260.

## Code Availability

The analysis pipeline and software used in this study are publicly available: analysis pipeline (xqtl-protocol, release 1.0.2; https://github.com/StatFunGen/xqtl-protocol); pecotmr (v.0.3.4; https://github.com/StatFunGen/pecotmr); and the cTWAS package for *M-cTWAS* (v.0.4.21; https://xinhe-lab.github.io/multigroup_ctwas/). The NIAGADS xQTL portal landing page provides the user-facing entry point to the FGMB resources, RWAS vignettes, and code to reproduce the figures of the article (https://adsp-fgc.niagads.org/xqtl-resources/fgmb_weights_database/).

## Acknowledgements

We thank the participants and investigators of the Alzheimer’s Disease Sequencing Project (ADSP) and the FunGen-AD consortium. This work was supported in part by NIH grants R35GM146868 (F.M.); R01AG076901, R01AG086467 and a grant from The Urbut Family Foundation (G.W.); R01AG93879, RF1AG077828, R21AG077168, and U01AG046170 (M.W.); U01AG072572 (P.L.D.); RF1AG058501 (C.C.); and R01AI175554 (X.H.).

## Author Contributions

G.W. and F.M. conceived, designed, and supervised the study. G.W. designed the statistical and bioinformatics pipeline. C.L., A.W., H.S. implemented the statistical and bioinformatics pipeline. C.L. performed data analysis and drafted the manuscript with input from G.W. and F.M. K.L., S.Q., and X.H. developed the *cTWAS* and *M-cTWAS* methods and analysis pipelines. Y.L. analyzed data. D.N. contributed to `pecotmr` R package software infrastructure. P.L.D., D.B., M.W., and C.C. provided data. All authors reviewed and edited the manuscript.

## Competing Interests

The authors declare no competing interests.

## Notes

### Competing Interest Statement

The authors have declared no competing interest.

### Author Declarations

Ethics committee/IRB of Columbia University gave ethical approval for this work.

### Summary of Updates

- Added funding information to the preamble. - Change format of the manuscript. - Fixed a few things.

