## Supplementary Information for "A Multi-Context Regulome-Wide Association Atlas for Genetic Studies of Aging Brain Disorders"

#### Supplementary Notes

##### S1 Molecular Dataset Composition and Extended Prediction Model Resources

*Molecular Dataset* refers to a source-specific molecular measurement panel used for predictor training, such as a brain region, cell type, or cell subtype measured for gene expression, protein abundance, or splicing, *i.e.*, CUIMC1, CUIMC2, or MIT. *Context* denotes the tissue, cell type, or cell subtype, whereas *Molecular Modality* denotes the type of molecular phenotype being modeled, *i.e.*, gene expression, protein abundance, or splicing. The *Molecular Trait* refer to a unique context–modality combination.

We constructed *cis*-genetic prediction models for gene expression, protein abundance, and splicing regulation across molecular datasets assembled by FunGen-AD, spanning ROSMAP[1], MSBB[2], and Knight-ADRC[3]. ROSMAP contributed bulk tissue RNA-seq from dorsolateral prefrontal cortex (DLPFC), posterior cingulate cortex (PCC), and anterior cingulate cortex (AC)[1]; DLPFC protein abundance[4]; bulk splicing models in DLPFC, PCC, and AC[5]; and three DLPFC single-nucleus RNA-seq resources. The single-nucleus resources included: (1) CUIMC1 from Fujita *et al.* 2024[6], with six major brain cell types — microglia (Mic), oligodendrocytes (Oli), oligodendrocyte progenitor cells (OPC), excitatory neurons (Exc), astrocytes (Ast), and inhibitory neurons (Inh) — and sample size of $N = 419$ for all cell types except OPC, for which $N = 418$; (2) the MIT dataset described by Xiong *et al.* 2023[7], with nine cell types and cell subtypes — Mic, Oli, OPC, Exc, Ast, Inh, astrocyte subtype 10 (Ast.10), microglia subtype 12 (Mic.12), and microglia subtype 13 (Mic.13) — and sample sizes of $N = 80$–387; and (3) the mega-analysis dataset, which integrates CUIMC1, MIT, and CUIMC2 single-nucleus RNA profiles for six major cell types (Mic, Oli, OPC, Exc, Ast, and Inh), with sample sizes of $N = 733$–737 across cell types. CUIMC2 is a recently generated ROSMAP DLPFC single-nucleus multiome (RNA + ATAC) dataset from approximately 240 individuals[8]; its RNA profiles contributed to the mega-analysis rather than being released as a separate FGMB prediction resource.

MSBB contributed four cortical RNA-seq datasets — frontal pole (FP, Brodmann area 10), superior temporal gyrus (STG, Brodmann area 22), parahippocampal gyrus (PHG, Brodmann area 36), and inferior frontal gyrus (IFG, Brodmann area 44) — with sample sizes of $N = 230$–274, as well as PHG protein abundance ($N = 184$).

Knight-ADRC contributed parietal cortex (PC) gene expression ($N = 354$) and protein abundance ($N = 412$). Sample sizes and imputable prediction model resources across molecular datasets are summarized in Table 1.

The model-building framework generalized beyond the 11 ROSMAP datasets used for the primary AD application. In ROSMAP bulk splicing, the eight-model framework identified 101,886 imputable productive (PR) or unproductive (UP) splice-site events across gene–molecular-trait pairs, comprising 11,049 unique imputable genes, with median cross-validation $R^2$ of 0.026–0.031 across contexts. Two additional Bayesian methods, including *SuSiE* and *mr.ash*, recovered 43,013 additional imputable splice events compared with *elastic-net* and *lasso* alone (Supplementary Figure S5). The framework also produced imputable predictors in additional ROSMAP single-nucleus resources, including MIT (12,307 genes tested, approximately 11,528 genes imputable in at least one context, 39,208 imputable gene–molecular-trait pairs, and median $R^2 \approx 0.06$ across nine cell types and cell subtypes) and the mega-analysis (27,428 imputable gene–molecular-trait pairs across six major cell types, with median $R^2 = 0.059$). In MSBB, the framework identified 42,143 imputable gene–molecular-trait pairs corresponding to 15,203 unique genes, with imputability rates of 80.8%–87.2% across datasets and 14,703 pairs recovered by the additional Bayesian or multivariate methods. In Knight-ADRC, it identified 9,303 imputable gene–molecular-trait pairs corresponding to 9,156 unique genes, with median $R^2 = 0.047$ and 2,803 pairs recovered by the additional Bayesian methods (Supplementary Figures S1–S4).

#### S2 Reusable Prediction and Fine-Mapping Protocol

FGMB is also designed as a protocol and software resource. The xqtl-protocol workflow provides a reproducible path from harmonized genotype and molecular data to prediction weights, RWAS association statistics, and downstream single-context *cTWAS* and multi-context *M-cTWAS* joint variant–gene fine-mapping outputs. The pecotmr R package supports model training, model selection, and integration of sparse-regression, Bayesian, and multivariate prediction methods. The cTWAS package provides the fine-mapping layer used to jointly analyze variants and predicted gene-level molecular traits. This protocol layer is part of the FGMB resource because it allows the same framework to be rerun as additional molecular datasets, cell subtypes, modalities, ancestries, or prediction methods become available.

Additional context-specific performance patterns supported the value of multivariate prediction. Multivariate methods such as *mr.mash* and *mvSuSiE* consistently showed higher imputability, particularly in molecular traits with smaller sample sizes or low cell proportions. For example, in DLPFC — a context with a larger sample size ($N = 777$) — the distribution of $R^2$ was relatively similar across all methods. By contrast, in PCC — a context with a smaller sample size ($N = 441$) — the multivariate models (*i.e.*, *mvSuSiE* and *mr.mash*) achieved higher $R^2$ than univariate methods (Figure 1D). A similar pattern appeared in cell-type contexts. The two multivariate methods showed higher $R^2$ distributions than univariate methods in Ast ($N = 419$), while performance was similar between univariate and multivariate methods in Exc ($N = 419$). Although Ast and Exc have the same sample size, their *effective* sample size differs because of cell-type proportions: Exc comprises 43% of the total cells, whereas Ast comprises only 13%[6]. This discrepancy reduces the effective sample size for Ast and lowers statistical power in univariate modeling. Multivariate models partially mitigated this limitation by leveraging shared effects across molecular traits to improve prediction where single-molecular-trait analyses are underpowered.

#### S3 Fine-Mapping Metrics and Credible Set Definitions

Single-group *cTWAS* and multi-context *M-cTWAS* perform Bayesian variant–gene fine-mapping analyses over candidate LD regions. The number of genes discovered by *cTWAS* and *M-cTWAS* are the result of a Bayesian multiple regression model within fine-mapped regions and are not directly comparable to the number of Bonferroni-significant RWAS genes, which result from a gene-by-gene (marginal) simple regression model. PIP and credible set criteria summarize posterior support within fine-mapped regions and are not directly comparable to marginal RWAS Bonferroni significance.

*M-cTWAS* produced a more selective set of prioritized signals. Across the fine-mapped regions, *M-cTWAS* identified 72 CSs, of which 35 were gene-driven (*i.e.*, the highest PIP within that CS was assigned to a gene) and 37 were SNP-driven (*i.e.*, the total PIP assigned to SNPs was greater than that assigned to genes within the CS[9]). In terms of composition, 29 *M-cTWAS* CSs contained only genes, 32 contained only SNPs, and 11 contained both genes and SNPs. In total, *M-cTWAS* supported 70 unique genes with suggestive evidence of non-zero effects (PIP $\geq$ 0.5 or inclusion in a CS). Among the 327 RWAS-significant gene–molecular-trait pairs, *M-cTWAS* prioritized 86 pairs, corresponding to 43 unique genes. By comparison, single-group *cTWAS* prioritized 146 RWAS-significant gene–molecular-trait pairs, corresponding to 63 unique genes, while 109 CSs were counted across all fine-mapped regions under the same PIP-or-CS criterion. This included 45 gene-driven and 64 SNP-driven CSs; 25 CSs contained only genes, 39 contained only SNPs, and 45 contained both genes and SNPs. Because a CS can contain multiple correlated genes, and because single-group *cTWAS* analyzes each molecular trait separately, this larger gene count should be interpreted as a less selective set of region-level candidates. Summary counts are provided in Supplementary Table S4.

#### S4 Functional Enrichment and WikiPathways Details

GO enrichment grouped the RWAS-significant genes into several AD-relevant biological themes. One theme involved immune and complement activity, including immunoglobulin-mediated immune response, B cell-mediated immunity, humoral immune response, and complement activation, with contributions from RWAS-significant genes such as *CR1*, *CR2*, *CLU*,*TREM2*, *FCER1G*,*BLNK*, and *NECTIN2*. Within this theme, several highlighted genes were novel, including *IGHG2*, *MPEG1*, *ZNF3*, and *RAB8B*. A second theme involved vesicle, endosomal, and membrane-associated biology, including intracellular vesicle, cytoplasmic vesicle, endocytic vesicle, vesicle-mediated transport, endosome, phagocytic vesicle, regulation of endocytosis, receptor-mediated endocytosis, membrane organization, protein localization to membrane, focal adhesion, cell-substrate junction, cell surface, receptor complex, and plasma membrane terms. These pathway-level terms were supported by RWAS-significant genes such as *APOE*, *PICALM*, *BIN1*, *CD2AP*, *ADAM10*, *TREM2*, and *RIN3*, and included several novel genes, such as *MPEG1*, *ZFYVE21*, *RAB8B*, *VGF*, *TMEM109*, *GP1BA*, *STX1B*, *ARHGAP27*, *PPP4C*, and *HIF3A*, and are consistent with AD biology involving cargo transport, endocytic uptake, receptor recycling, membrane remodeling, *APP* processing, lipid handling, and A$\beta$ clearance[10–16]. A third theme involved amyloid and lipid handling and glial response, including amyloid-beta metabolic process, astrocyte activation, and lipoprotein particle clearance terms, supported by RWAS-significant genes such as *APOE*, *APOC1*, *APOC2*, *APOC4-APOC2*, *APP*, *ADAM10*, *CLU*, *TREM2*, and *GRN*.

The same enrichment query also identified WikiPathways gene sets corresponding to annotated copy number variant (CNV) regions: 16p11.2 proximal deletion and 10q22–q23 copy number variation. These overlaps primarily reflect clustering of RWAS-significant genes within annotated genomic regions. The 16p11.2 overlap included several genes with dosage-sensitive neurodevelopmental relevance, whereas *ADAM10* in the 10q22–q23 set has stronger prior AD support as an AD risk locus involved in *APP* metabolism[17]. In contrast, genes such as *ASPHD1*, *MAT1A*, and *DYDC1* currently provide limited AD-specific mechanistic evidence. These pathway-level results are therefore best interpreted as regional functional context rather than structural-variant involvement or locus-level fine-mapping evidence.

### Supplemental Figures

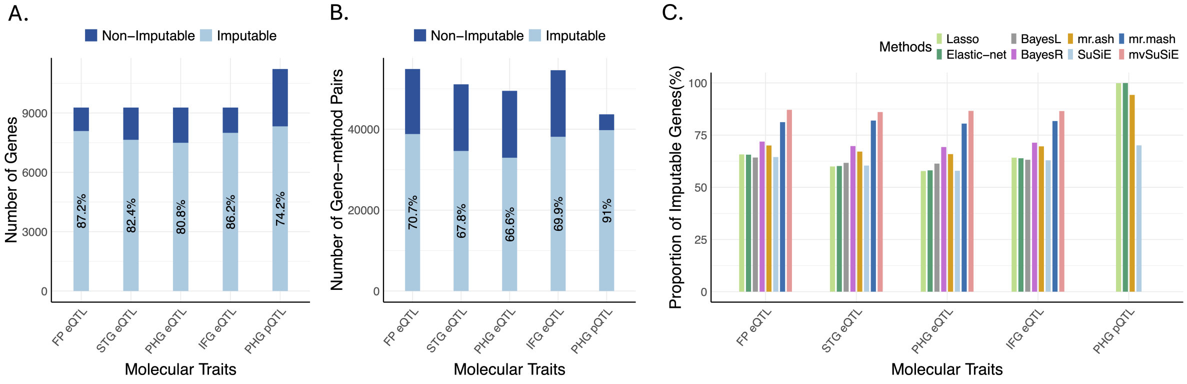


**Figure S1. MSBB RWAS weights.**

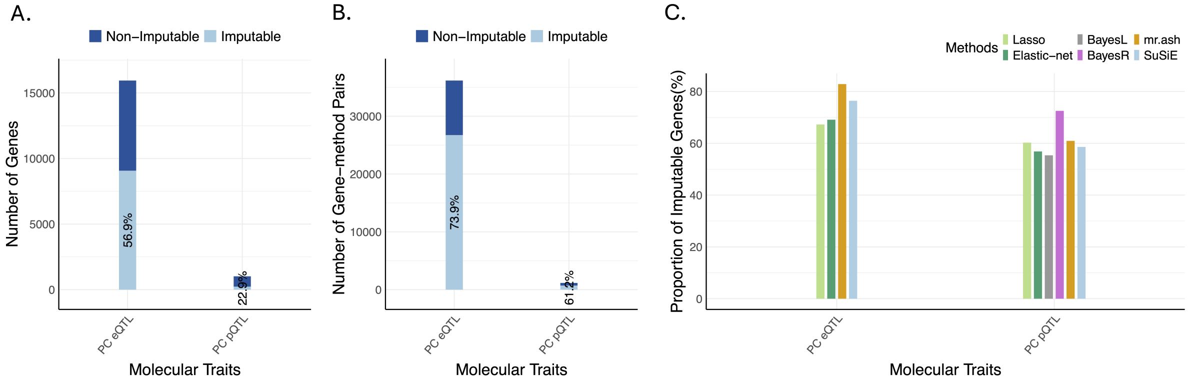


**Figure S2. Knight-ADRC[3] RWAS weights.**

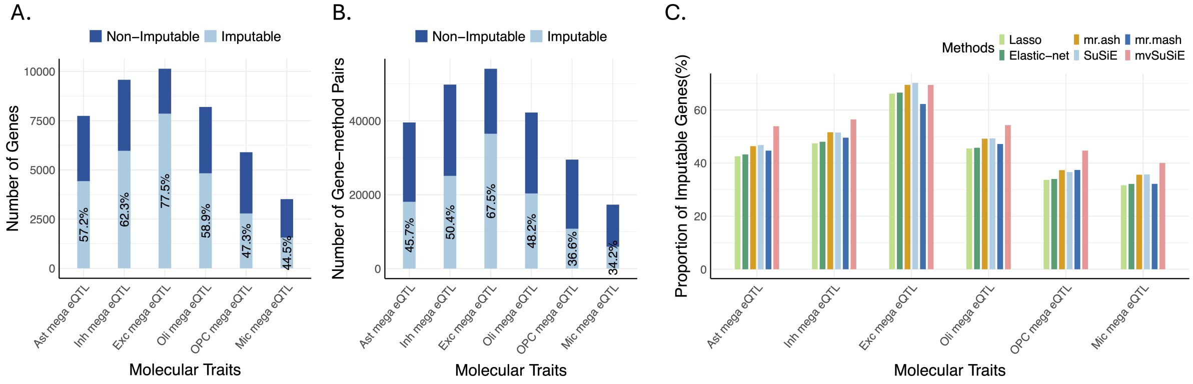


**Figure S3. ROSMAP mega-analysis[8] RWAS weights.**
ROSMAP RWAS weights derived from pooled single-nucleus RNA-seq eQTL data across ROSMAP CUIMC1[6], MIT[7], and the recently generated CUIMC2 single-nucleus multiome (RNA + ATAC) dataset[8].

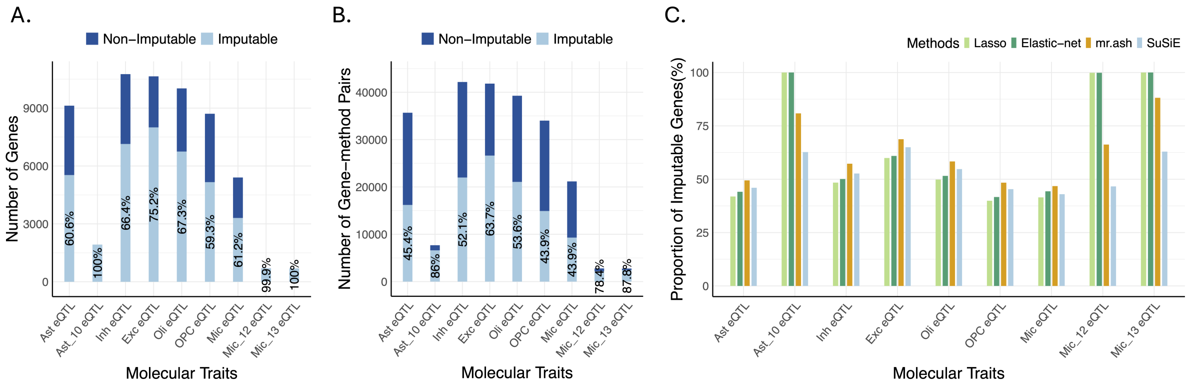

**Figure S4. ROSMAP MIT[7] RWAS weights.**

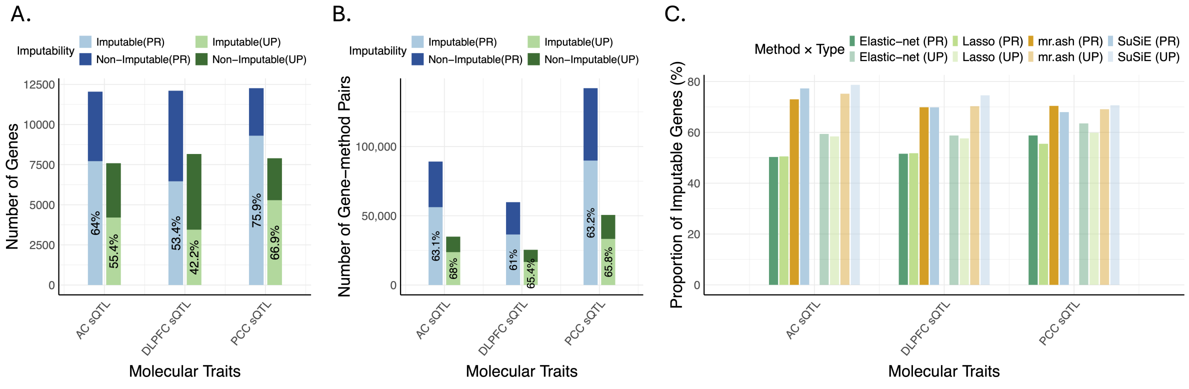

**Figure S5. ROSMAP splicing QTL[5] RWAS weights.**
Splicing events were annotated based on predicted coding consequences as productive (PR), unproductive (UP), no-effect (NE), or indeterminate (IN). Only productive and unproductive events were used for RWAS analyses.

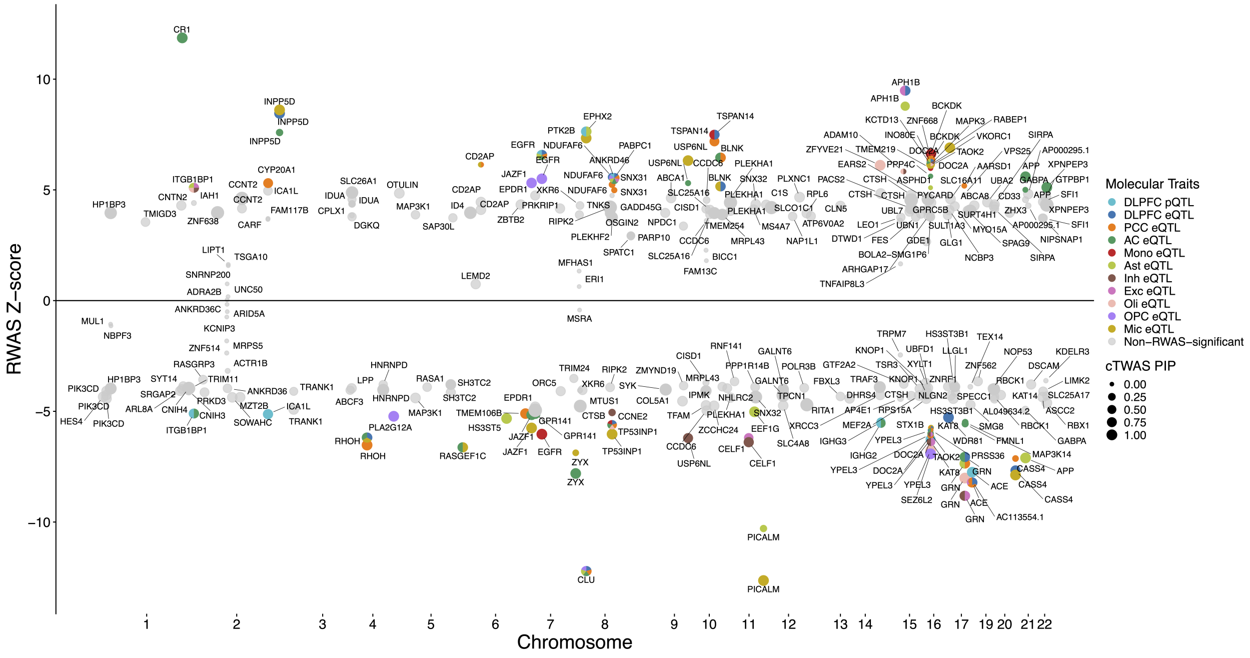


**Figure S6. Distribution of RWAS z-scores for single-group *cTWAS*-prioritized gene–molecular-trait pairs.**

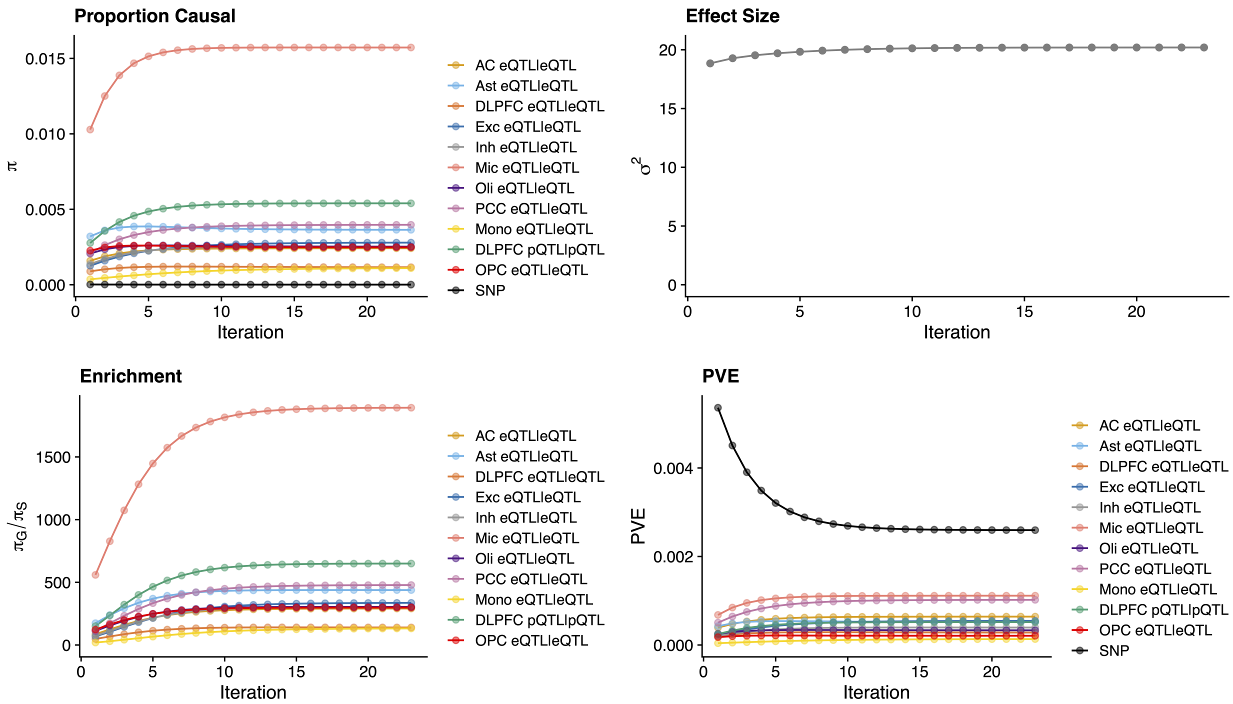


**Figure S7. *M-cTWAS* global prior convergence plot.**
*M-cTWAS* global parameter estimation convergence plot shows estimated prior inclusion probability for each group across iterations (Proportion Causal), estimated prior effect-size variance across iterations (Effect Size), enrichment relative to SNPs (Enrichment), and proportion of variance explained (PVE).

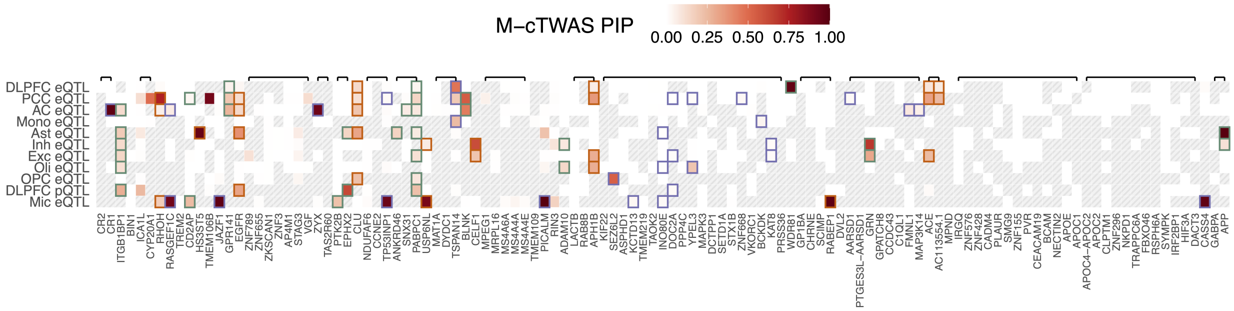


**Figure S8.** ***M-cTWAS*** **PIP distribution across genes that were RWAS-significant in at least one molecular trait.**
This heatmap shows the distribution of *M-cTWAS* posterior inclusion probabilities (PIP; range 0–1) across gene–molecular-trait pairs for genes that were RWAS-significant in at least one molecular trait. Each box corresponds to a gene–molecular-trait pair. Striped gray boxes denote gene–molecular-trait pairs for which the expression prediction model did not satisfy the imputability criteria from any modeling method or did not have valid RWAS results. Outlined boxes alternating purple, green, and orange indicate gene–molecular-trait pairs that were included in a credible set (CS), while brackets above the heatmap indicate genes belonging to the same LD region block.

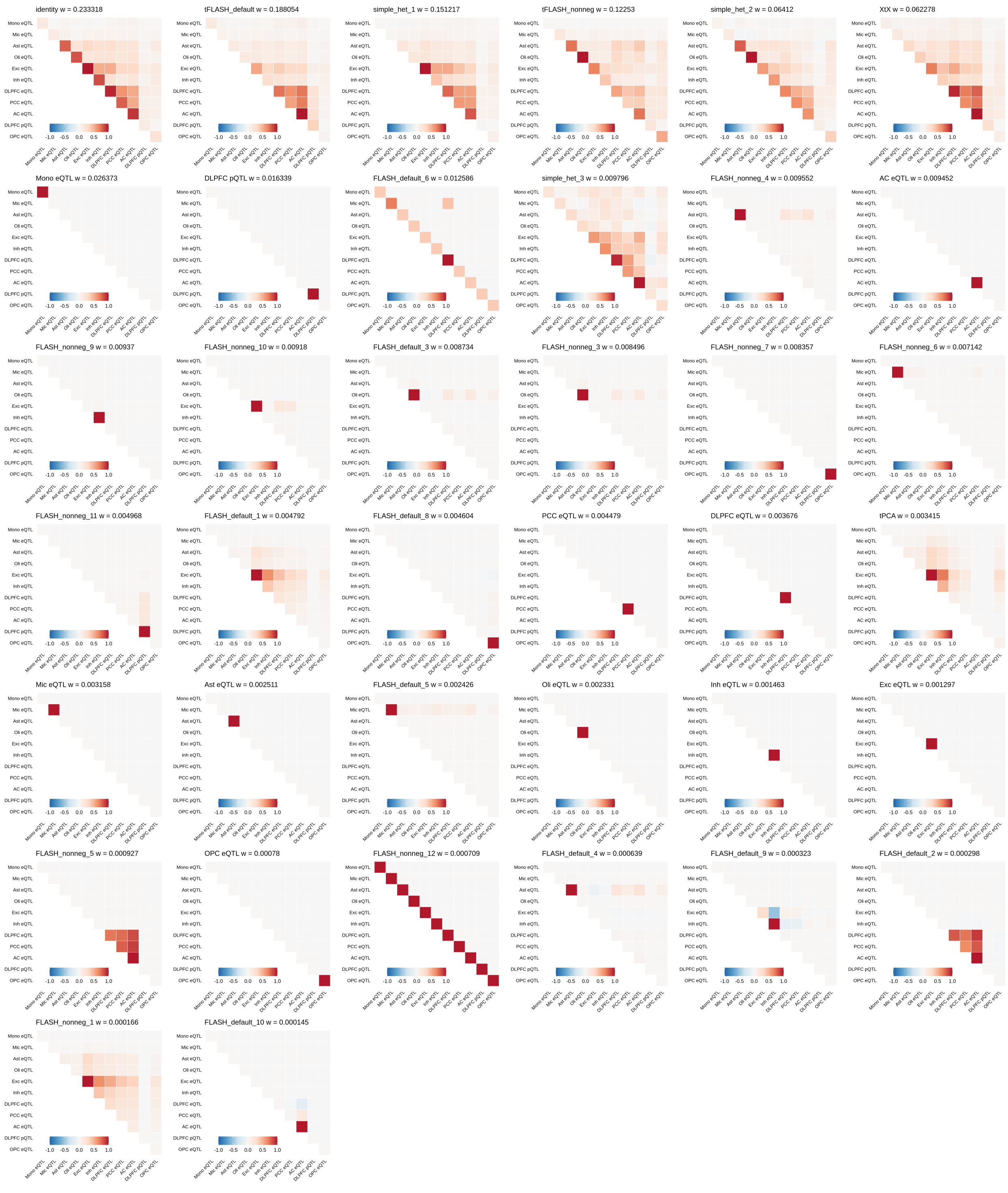


**Figure S9. Mixture prior components used for multivariate expression predictor training in ROSMAP.**
These covariance matrices represent patterns of shared genetic effects across molecular traits, with component weights shown on top of each plot, estimated by *mr.mash* after initialization from the *Extreme Deconvolution* framework. Components with negligible weights ($w_0 \leq 1 \times 10^{-4}$) were excluded prior to multivariate model fitting (see Methods, Training Molecular Trait Predictors for Multi-Context, Multi-Omics Data).

### Supplemental Tables

| Gene Name | Molecular Trait | Method | Cross Validation $R^2$ | RWAS *p*-value |
|---|---|---|---|---|
| *NECTIN2* | Ast eQTL | *SuSiE* | 0.02563 | $2.09 \times 10^{-31}$ |
| *CLPTM1* | DLPFC eQTL | *SuSiE* | 0.02027 | $5.62 \times 10^{-21}$ |
| *HIF3A* | PCC eQTL | *SuSiE* | 0.01164 | $7.74 \times 10^{-21}$ |
| *ZNF576* | Ast eQTL | *SuSiE* | 0.01814 | $1.76 \times 10^{-20}$ |
| *APOE* | Mic eQTL | *mvSuSiE* | 0.10442 | $1.23 \times 10^{-19}$ |
| *RSPH6A* | PCC eQTL | *mr.mash* | 0.06487 | $2.60 \times 10^{-18}$ |
| *BCAM* | AC eQTL | *elastic net* | 0.01837 | $6.17 \times 10^{-15}$ |
| *PTK2B* | Mic eQTL | *mr.ash* | 0.24346 | $2.14 \times 10^{-13}$ |
| *ZNF428* | Oli eQTL | *elastic net* | 0.01045 | $7.09 \times 10^{-13}$ |
| *ZNF155* | Mono eQTL | *SuSiE* | 0.04498 | $8.80 \times 10^{-12}$ |
| *FBXO46* | Ast eQTL | *BayesR* | 0.01879 | $1.05 \times 10^{-11}$ |
| *SMG9* | Mic eQTL | *BayesL* | 0.02359 | $1.56 \times 10^{-11}$ |
| *CR2* | DLPFC eQTL | *BayesL* | 0.03321 | $2.17 \times 10^{-11}$ |
| *PTGES3L-AARSD1* | PCC eQTL | *BayesL* | 0.03094 | $7.30 \times 10^{-11}$ |
| *TAS2R60* | Mono eQTL | *lasso* | 0.06535 | $1.15 \times 10^{-10}$ |
| *VGF* | PCC eQTL | *BayesR* | 0.01470 | $2.06 \times 10^{-10}$ |
| *ZNF668* | PCC eQTL | *mr.ash* | 0.10858 | $2.08 \times 10^{-10}$ |
| *MPND* | Ast eQTL | *elastic net* | 0.01511 | $2.92 \times 10^{-10}$ |
| *DYDC1* | AC eQTL | *SuSiE* | 0.06957 | $6.34 \times 10^{-10}$ |
| *MAPK3* | AC eQTL | *SuSiE* | 0.05439 | $7.38 \times 10^{-10}$ |
| *KCTD13* | Mic eQTL | *mr.ash* | 0.01717 | $7.69 \times 10^{-10}$ |
| *VKORC1* | AC eQTL | *elastic net* | 0.04323 | $8.15 \times 10^{-10}$ |
| *AP4M1* | Inh eQTL | *SuSiE* | 0.09028 | $9.44 \times 10^{-10}$ |
| *ZFYVE21* | Oli eQTL | *mr.ash* | 0.01143 | $9.79 \times 10^{-10}$ |
| *NSF* | Mono eQTL | *mvSuSiE* | 0.02703 | $1.22 \times 10^{-9}$ |
| *CADM4* | Ast eQTL | *elastic net* | 0.01052 | $2.13 \times 10^{-9}$ |
| *MPEG1* | PCC eQTL | *BayesR* | 0.01261 | $2.25 \times 10^{-9}$ |
| *TMEM219* | Mono eQTL | *mr.mash* | 0.01854 | $2.28 \times 10^{-9}$ |
| *SCIMP* | Mic eQTL | *mvSuSiE* | 0.13653 | $2.94 \times 10^{-9}$ |
| *GPATCH8* | Ast eQTL | *mr.ash* | 0.01093 | $3.66 \times 10^{-9}$ |
| *CCDC43* | Ast eQTL | *BayesL* | 0.02028 | $9.12 \times 10^{-9}$ |
| *TRAPPC6A* | PCC eQTL | *BayesR* | 0.01628 | $9.82 \times 10^{-9}$ |
| *C1QL1* | DLPFC eQTL | *BayesL* | 0.01232 | $1.10 \times 10^{-8}$ |
| *AC005670.2* | PCC eQTL | *mvSuSiE* | 0.06216 | $1.15 \times 10^{-8}$ |
| *SYMPK* | DLPFC pQTL | *BayesL* | 0.01482 | $1.60 \times 10^{-8}$ |
| *MAT1A* | AC eQTL | *mvSuSiE* | 0.06557 | $1.69 \times 10^{-8}$ |
| *IRGQ* | Oli eQTL | *BayesL* | 0.01218 | $2.00 \times 10^{-8}$ |
| *FMNL1* | AC eQTL | *mr.ash* | 0.01959 | $2.61 \times 10^{-8}$ |
| *ANKRD46* | Ast eQTL | *mvSuSiE* | 0.03901 | $3.05 \times 10^{-8}$ |
| *MAP3K14* | AC eQTL | *SuSiE* | 0.02670 | $3.98 \times 10^{-8}$ |
| *IGHG3* | DLPFC pQTL | *elastic net* | 0.22824 | $4.16 \times 10^{-8}$ |
| *STH* | DLPFC eQTL | *mvSuSiE* | 0.02513 | $4.49 \times 10^{-8}$ |
| *ZNF3* | Ast eQTL | *mr.mash* | 0.01746 | $5.53 \times 10^{-8}$ |
| *SETD1A* | DLPFC eQTL | *mr.mash* | 0.01396 | $6.74 \times 10^{-8}$ |
| *MRPL16* | Exc eQTL | *mr.ash* | 0.01927 | $7.20 \times 10^{-8}$ |
| *TMEM109* | Mono eQTL | *BayesL* | 0.01949 | $7.28 \times 10^{-8}$ |
| *GP1BA* | DLPFC eQTL | *SuSiE* | 0.02212 | $9.74 \times 10^{-8}$ |
| *HS3ST5* | Ast eQTL | *SuSiE* | 0.13381 | $1.01 \times 10^{-7}$ |
| *DVL2* | DLPFC pQTL | *SuSiE* | 0.01775 | $1.08 \times 10^{-7}$ |
| *CYP20A1* | PCC eQTL | *mr.ash* | 0.01196 | $1.12 \times 10^{-7}$ |
| *WDR81* | DLPFC eQTL | *mvSuSiE* | 0.02234 | $1.34 \times 10^{-7}$ |
| *PLEKHM1* | AC eQTL | *BayesR* | 0.39425 | $1.62 \times 10^{-7}$ |
| *PLA2G12A* | OPC eQTL | *BayesR* | 0.01215 | $1.75 \times 10^{-7}$ |
| *AARSD1* | PCC eQTL | *mvSuSiE* | 0.02314 | $2.14 \times 10^{-7}$ |
| *ZNF789* | Exc eQTL | *lasso* | 0.02307 | $2.54 \times 10^{-7}$ |
| *ZNF655* | Mono eQTL | *mr.mash* | 0.02075 | $2.65 \times 10^{-7}$ |
| *DCTPP1* | Mono eQTL | *mr.mash* | 0.02298 | $2.87 \times 10^{-7}$ |
| *XPNPEP3* | AC eQTL | *SuSiE* | 0.03761 | $3.15 \times 10^{-7}$ |
| *GPR141* | AC eQTL | *SuSiE* | 0.14215 | $3.34 \times 10^{-7}$ |
| *TMEM106B* | PCC eQTL | *mr.mash* | 0.01995 | $3.34 \times 10^{-7}$ |
| *PLAUR* | Mono eQTL | *SuSiE* | 0.02372 | $3.54 \times 10^{-7}$ |
| *NKPD1* | DLPFC eQTL | *SuSiE* | 0.01032 | $4.19 \times 10^{-7}$ |
| *CCNE2* | Inh eQTL | *mr.ash* | 0.01391 | $4.34 \times 10^{-7}$ |
| *DACT3* | Ast eQTL | *mr.mash* | 0.02683 | $4.45 \times 10^{-7}$ |
| *KIF22* | Oli eQTL | *mr.ash* | 0.02031 | $4.61 \times 10^{-7}$ |
| *RAB8B* | DLPFC pQTL | *mvSuSiE* | 0.06097 | $4.96 \times 10^{-7}$ |
| *EEF1G* | Ast eQTL | *BayesR* | 0.01485 | $5.12 \times 10^{-7}$ |
| *GABPA* | AC eQTL | *mr.ash* | 0.26374 | $5.63 \times 10^{-7}$ |
| *ZNF652* | AC eQTL | *SuSiE* | 0.04839 | $5.92 \times 10^{-7}$ |

**Table S1. Molecular-Trait-specific RWAS associations.**

| GWAS cohort | *N* case | *N* control |
|---|---|---|
| Bellenguez_2022[18] | 111,326 | 677,663 |
| Bellenguez_EADB_2022[18,19] | 20,301 | 21,839 |
| Bellenguez_EADI_2022[18,20] | 2,400 | 6,338 |
| Bellenguez_GRACE_2022[18,21] | 6,497 | 6,785 |
| Jansen_2021[22] | 71,880 | 383,378 |
| Kunkle_Stage1_2019[17] | 21,982 | 41,944 |
| Wightman_Full_2021[23] | 90,338 | 1,036,225 |
| Wightman_Excluding23andMe_2021[23] | 86,531 | 676,386 |
| Wightman_ExcludingUKBand23andME_2021[23] | 39,918 | 35,814 |

**Table S2. Sample size GWAS cohorts.**

| Gene Name | Replicated Molecular Trait(s) Replicated Across All 4 GWAS Studies | Molecular Trait(s) Replicated in At Least 2 GWAS Studies |
|---|---|---|
| *APOC1* | AC eQTL[*,†,§,‡]; DLPFC eQTL[*,†,§,‡] | DLPFC pQTL[*,‡]; Mic eQTL[*,†] |
| *APOC2* | AC eQTL[*,†,§,‡]; DLPFC eQTL[*,†,§,‡]; PCC eQTL[*,†,§,‡] | |
| *APOC4-APOC2* | AC eQTL[*,†,§,‡]; PCC eQTL[*,†,§,‡] | |
| *APOE* | Mic eQTL[*,†,§,‡] | |
| *BCAM* | AC eQTL[*,†,§,‡] | |
| *BIN1* | Exc eQTL[*,†,§,‡]; Mic eQTL[*,†,§,‡] | AC eQTL[*]; DLPFC pQTL[*,§,‡]; DLPFC eQTL[*]; PCC eQTL[*] |
| *CADM4* | Ast eQTL[*,†,§,‡] | |
| *CD2AP* | Mic eQTL[*,†,§,‡]; PCC eQTL[*,†,§,‡] | Exc eQTL[*,‡] |
| *CEACAM19* | AC eQTL[*,†,§,‡]; DLPFC eQTL[*,†,§,‡]; Exc eQTL[*,†,§,‡]; Inh eQTL[*,†,§,‡]; PCC eQTL[*,†,§,‡] | |
| *CLPTM1* | DLPFC eQTL[*,†,§,‡] | Oli eQTL[†,§]; AC eQTL[‡]; Mic eQTL[‡]; PCC eQTL[‡]; Monocyte eQTL[‡] |
| *CLU* | AC eQTL[*,†,§,‡]; Ast eQTL[*,†,§,‡]; DLPFC eQTL[*,†,§,‡]; OPC eQTL[*,†,§,‡]; PCC eQTL[*,†,§,‡] | |
| *DACT3* | Ast eQTL[*,†,§,‡] | |
| *FBXO46* | Ast eQTL[*,†,§,‡] | |
| *HIF3A* | PCC eQTL[*,†,§,‡] | |
| *INPP5D* | DLPFC eQTL[*,†,§,‡]; Mic eQTL[*,†,§,‡] | AC eQTL[*,§,‡] |
| *IRF2BP1* | DLPFC pQTL[*,†,§,‡] | Oli eQTL[*,†,§] |
| *IRGQ* | | Oli eQTL[*]; AC eQTL[†,§,‡] |
| *MS4A4A* | DLPFC eQTL[*,†,§,‡]; Mic eQTL[*,†,§,‡]; PCC eQTL[*,†,§,‡] | AC eQTL[*,‡] |
| *MS4A4E* | AC eQTL[*,†,§,‡]; DLPFC eQTL[*,†,§,‡] | Mic eQTL[*,‡] |
| *MS4A6A* | DLPFC eQTL[*,†,§,‡]; Mic eQTL[*,†,§,‡] | |
| *NECTIN2* | Ast eQTL[*,†,§,‡] | |
| *PICALM* | Ast eQTL[*,†,§,‡]; Mic eQTL[*,†,§,‡]; Monocyte eQTL[*,†,§,‡] | Oli eQTL[*,‡] |
| *PTK2B* | Mic eQTL[*,†,§,‡] | |
| *PVR* | AC eQTL[*,†,§,‡]; DLPFC eQTL[*,†,§,‡] | DLPFC pQTL[*,§,‡]; Exc eQTL[†,§,‡]; PCC eQTL[§,‡] |
| *RSPH6A* | PCC eQTL[*,†,§,‡] | |
| *TRAPPC6A* | PCC eQTL[*,†,§,‡] | |
| *USP6NL* | Mic eQTL[*,†,§,‡] | AC eQTL[*,†,‡]; Inh eQTL[*,‡] |
| *ZNF296* | DLPFC eQTL[*,†,§,‡]; PCC eQTL[*,†,§,‡] | |
| *ZNF428* | Oli eQTL[*,†,§,‡] | |
| *ZNF576* | Ast eQTL[*,†,§,‡] | |

**Table S3. Cross-GWAS RWAS significant genes.**
This table includes only genes with at least one molecular trait replicated across all four GWAS studies[*,†,§,‡].
[*] Bellenguez et al. 2022[18];
[†] Kunkle et al. 2019[17];
[§] Jansen et al. 2021[22];
[‡] Wightman et al. 2021[23].

| | Prioritized Gene–molecular-trait Pairs | CS | Gene-driven CS | SNP-driven CS | Gene-only CS | SNP-only CS |
|---|---|---|---|---|---|---|
| *M-cTWAS* | 86 | 72 | 35 | 37 | 29 | 32 |
| *cTWAS* | 146 | 109 | 45 | 64 | 25 | 39 |

**Table S4. Summary of *cTWAS* and *M-cTWAS* Fine-Mapping Results.**
Single-group *cTWAS* and multi-context *M-cTWAS* were run as Bayesian joint variant–gene fine-mapping analyses over candidate LD regions. PIP and credible set criteria summarize posterior support within fine-mapped regions and are not directly equivalent to marginal RWAS Bonferroni significance. The posterior prioritization on the molecular-trait pair focus on the subset overlapping the 327 RWAS-significant gene–molecular-trait pairs. We further summarized credible set (CS) composition according to whether posterior support was primarily assigned to gene-level predictors or SNP-level effects. SNP-driven CSs were those in which the sum of PIP assigned to SNPs exceeded the sum of PIP assigned to gene-level predictors, whereas gene-driven CSs were those in which the sum of PIP assigned to gene-level predictors exceeded the sum PIP assigned to SNPs[9].
